# Complexity of Infection and *Plasmodium falciparum* Circumsporozoite Protein Diversity Prior to Malaria Vaccine Implementation in Kaelé Health District, Cameroon, 2022-2023

**DOI:** 10.64898/2026.01.17.26344325

**Authors:** Innocent M. Ali, Brenda Vasquez Martinez, Valery P. K. Tchuenkam, Jacob M. Sadler, Catherine C. Gorman, Sandrine E. Nsango, Voundi Voundi Junior, Ateba Joel Marcellin, Grace Yimga Wanda, Jeffrey A. Bailey, Rhoel R. Dinglasan, Jessica T. Lin, Jonathan J. Juliano

## Abstract

Malaria remains a major public health concern in Cameroon, with *Plasmodium falciparum* responsible for most morbidity and mortality, particularly among children under five. In response to rising cases, Cameroon began implementing the RTS,S/AS01 malaria vaccine in early 2024. Given the vaccine’s strain-specific efficacy, understanding antigenic diversity and complexity of infection (COI) is critical for evaluating long-term impact. We analyzed 100 *P. falciparum*–positive dried blood spots collected in Mapoussere, Kaele Health District (2022–2023). Using the 4CAST amplicon sequencing assay, we targeted four genes: *csp* (circumsporozoite protein), *ama* (apical membrane antigen 1), *sera2* (serine repeat antigen 2), and *trap* (thrombospondin-related anonymous protein). Haplotypes were identified using SeekDeep, and diversity metrics including heterozygosity (He), nucleotide diversity (π), and selection statistics (Tajima’s D, Fu and Li’s D*, F*) were computed. We successfully genotyped *csp* in 35% of samples, identifying 22 haplotypes (He = 0.908; π = 0.021). The vaccine-matched haplotype was present in 20% of genotyped infections. The T cell TH2 and TH3 epitopes of *csp* showed signs of balancing selection. Both *ama* and *sera2* exhibited higher genotyping success and diversity, with *ama* showing significant Tajima’s D values. COI was highest for *ama* (mean COI = 2.8), followed by *sera2* (2.1), *csp* (1.3), and *trap* (1.2). This study provides a baseline of *P. falciparum* antigenic diversity and COI in a vaccine-targeted region. The presence of vaccine-matched strains and high diversity in TH2/TH3 epitopes in *csp* may influence vaccine efficacy. Continued molecular surveillance is essential to monitor antigenic shifts and guide future strategies.

## INTRODUCTION

Malaria remains an important public health problem in Cameroon, with the entire country being endemic for malaria transmission, and ranking 11th on the list of countries with the most malaria cases.(National Malaria Control Program, Cameroon, n.d.) In 2023, there were over 7.3 million cases of malaria in Cameroon, with an estimated 11,300 deaths.(World Health Organization, 2024) The majority of morbidity and mortality occurs in children under 5 years of age and is due to *Plasmodium falciparum*. Cameroon implements major interventions for controlling malaria, including treatment with effective artemisinin combination therapies (ACT), the use of long-lasting insecticide-treated bednets (LLIN), and indoor residual spraying (IRS). To date, validated and candidate mutations associated with artemisinin partial resistance have not been documented. Despite effective therapy being available, the number of cases increased by nearly 1.2 million per year between 2019 and 2023.(World Health Organization, 2024) In response, Cameroon was one of the first countries to roll out RTS,S/ASO1(Mosquirix™), which began in January 2024, across 42 priority health districts, with the goal of countrywide coverage by the end of 2026. (Ndoula et al., 2024) Cameroon’s Expanded Program on Immunization (EPI) reported that districts included in the initial rollout of the RTS,S malaria vaccine experienced a 17% greater reduction in all-cause hospitalization and outpatient clinic visits for children under five, compared to districts without the vaccine.(ReliefWeb, n.d.) Additionally, around 60% of vaccinated districts saw a decline in overall under-five mortality, while 57% recorded fewer malaria-related deaths among children in this age group.(ReliefWeb, n.d.)

RTS,S/ASO1 has been associated with a 55.1% (95% CI 50.5 to 59.3) reduction in clinical malaria in children aged 5-17 months during the 12 months of follow-up post-immunization, and 8·3% (95% CI 23·3 to 32·9) over 4 years, however, there is evidence of strain-specific efficacy, with lower efficacy against heterologous strains, defined by C-terminus of the *P. falciparum* circumsporozoite protein (*csp*) containing two critical T cell epitopes, TH2 and TH3. (RTS,S Clinical Trials Partnership et al., 2011; Neafsey et al., 2015; RTS,S Clinical Trials Partnership, 2015) Sequencing of *P. falciparum* circumsporozoite protein (*csp*), the gene encoding the antigen in the vaccine, shows differences in parasite populations by continent (Barry et al., 2009), but relatively few differences between populations within a continent.(Aragam et al., 2013) This is potentially due to convergent evolution of the antigen due to interaction with the human immune system, leading the gene to beunder balancing selection. Tracking of *csp* variants in the population can be useful in understanding whether selection pressure of the vaccine on the parasite population results in vaccine escape, whereby non-vaccine-matched genetic variants have a selective advantage and increase in frequency over time. To date, there is little information on *csp* diversity in Cameroon. One study reported 25 distinct haplotypes in the C-terminus of *csp* among 57 successfully sequenced samples from different areas of Cameroon collected in 2019.(Kojom Foko et al., 2024) A second study did not report haplotypes, but found 51 single nucleotide polymorphisms in *csp* from a subset of 117 parasitemic samples collected in 2024, primarily in the TH2 and TH3 epitopes.(Efeti et al., 2025)

*P. falciparum* infections in Africa commonly contain more than one strain of malaria, and thus infections may contain strains representing more than one *csp* variant based on the TH2 and TH3 variation. Complexity of infection (COI), a measure of the number of strains or genetic variants found within a single individual or parasite isolate, is loosely correlated to transmission intensity, (Watson et al., 2021) and therefore varies by geographic region. Within Cameroon, COI has been evaluated by amplicon sequencing (Sadler et al., 2024), microsatellite analysis (Atuh et al., 2022; Biabi et al., 2023), and fragment length polymorphism analysis.(Wanji et al., 2012; Biabi et al., 2023) Multi-strain infections were common across these studies.

As part of an effort to support the rollout and evaluation of RTS,S vaccine effectiveness in Cameroon, we assessed the diversity of the C-terminal TH2 and TH3 epitopes and flanking regions, as well as the COI for 100 *P. falciparum* positive samples collected in the Kaelé health district in the Far North region of Cameroon during December 2022. This area is one of the original 42 districts where RTS,S/ASO1 was rolled out, providing an opportunity to assess baseline malaria diversity for the district. We leveraged a published multiplex amplicon sequencing assay, 4CAST, which targets variable regions of *P. falciparum csp*, apical membrane antigen 1 (*ama*), serine repeat antigen 2 (*sera2*), and thrombospondin-related anonymous protein (*trap*).(LaVerriere et al., 2022)

## METHODS

### Ethics Statement

Ethical clearance for this study was obtained from the National Ethics Committee for Human Health Research under approval No 2021/12/1428/CE/CNERSH/SP on the 22nd of December, 2021. De-identified dried blood spots (DBS) were brought to the University of North Carolina at Chapel Hill for genotyping and are considered non-human subjects research (24-0777). Approved consent forms were used to obtain assent and/or parental authorization before initiating any data or blood collection.

### Participant samples

Sample and data collection were conducted in participants’ households, targeting asymptomatic children aged 1 to 14 years as part of a malaria survey conducted in Kaelé health district in the Far North region of Cameroon during December 2022. Trained research technicians administered consent forms in English, French, or Fufulde. Following written consent or parental authorization, fingerstick blood samples were obtained, and hemostasis was ensured using sterile bandages. Approximately 250 µL of blood was collected via capillary tube; 5–6 µL was immediately used for a malaria rapid diagnostic test, and upon a positive result, two spots of roughly 80-100 µL each were applied onto Whatman No. 3 filter paper. The filter papers were dried overnight in a shaded, fly-free environment, carefully folded, stored in ziplock bags with silica gel desiccants, and transported to the laboratory. Samples were maintained at room temperature until DNA extraction was performed.

### Targeted amplicon sequencing

Each DBS was punched into 96-well plates, and DNA was extracted from 3 × 6mm punches using a Chelex-Tween extraction method (dx.doi.org/10.17504/protocols.io.kxygx414kl8j/v1 and dx.doi.org/10.17504/protocols.io.dm6gpm5mjgzp/v1). Multiplex amplicon sequencing of the *csp, ama, sera2*, and *trap* genes was conducted using 4CAST.(LaVerriere et al., 2022) Details of the application of this method are provided at protocols.io (dx.doi.org/10.17504/protocols.io.eq2ly4qdmlx9/v1). Amplicons were pooled and sequenced as part of a 2X300bp MiSeq i100 25M run at the UNC High Throughput Sequencing Facility.

## Data analysis

Demultiplexed amplicons were analyzed using SeekDeep (Hathaway et al., 2018) with a minimum within-sample haplotype cluster size of 500 reads and a minimum within-sample allele frequency cut-off of 5%, applying whichever was most restrictive per sample. Cutoffs were determined using sequence data from controls, including mocked DBS of *P. falciparum* 3D7 parasites (MRA-102) at different parasitemias, (described elsewhere (Gaither et al., 2025)) and 2 mocked mixtures of genomic DNA (MRA-102G for *P. falciparum* strain 3D7 and MRA-152G for strain 7G8) (Mix 1: 3D7 80% and 7G8 20%; Mix 2: 7G8 95% and 3D7 5%).

Statistical analysis and graphing were completed with Graphpad Prism (v10.5.0) (Graphpad Software, Boston, MA). Heterozygosity (He) for each amplicon, which is the probability that two amplicon haplotypes will be different, was determined as 1-(p1^2^+p2^2^….+pn^2^), where p1, p2, etc. are the population frequencies of each haplotype and n is the total number of haplotypes in the population. Sequence logos were made using https://weblogo.berkeley.edu/logo.cgi. Median-joining phylogeny was generated using MEGA (v11).

Sequence analysis was carried out using DnaSP v.6.(Rozas et al., 2017) Nucleotide diversity (π) was used to quantify genetic variation within populations, where π represents the average number of nucleotide differences per site between all possible pairs of DNA sequences in a sample. To calculate π, aligned sequences were analyzed by computing the number of nucleotide differences between each pair, dividing by the sequence length to obtain per-site differences, and averaging across all unique sequence pairs. The formula is:

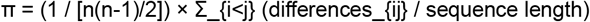

where n is the number of sequences. To detect deviations from neutral evolution, three standardized statistics were computed: Tajima’s D, Fu and Li’s D*, and Fu and Li’s F*. These metrics compare different estimates of genetic diversity to infer selection or demographic history:

1. Tajima’s D compares π to Watterson’s θ (θ□), which is based on the number of segregating sites. Under neutrality and constant population size, π ≈ θ□ and D ≈ 0. Deviations were interpreted as follows:
  – D < –2: Suggests an excess of rare alleles, consistent with purifying selection or population expansion.
  – D > +2: Indicates an excess of intermediate-frequency alleles, consistent with balancing selection or population bottlenecks.
2. Fu and Li’s D* compares the number of segregating sites (S) to the number of singletons (η_1_), using the simplified formula:

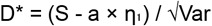
3. Fu and Li’s F* compares π to η_1_:

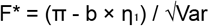

where a and b are constants based on the sample size. For both D* and F*, values < –2 indicate an excess of singletons (suggesting recent expansion or purifying selection), while values > +2 suggest a deficit of singletons (consistent with balancing selection or population structure). These statistics were applied to each locus as a complete haplotype, as well as using a sliding window (window: 25bp, slide: 5bp).

## RESULTS

### Participant characteristics

Participants were recruited directly from their homes, with a maximum of two children enrolled per household. This study included 100 samples that tested highly positive for *P. falciparum* by microscopy, with a geometric mean parasite density of 5744.12 parasites/µL (95%CI: 3965.51-7522.73). Participants ranged in age from 3 months to 14 years. The most represented age group was 3 to 59 months, accounting for 60% of the samples, while children aged 6 to 14 years made up the remaining 40%. In terms of gender distribution, males comprised 55% of the participants, and females 45%.

### Sequencing quality and success

Results of sequencing controls are shown in **Supplemental Table 1**. Overall, *ama* and *sera2* performed the most robustly across samples, providing similar within-sample allele frequency estimates in the mixtures and most samples with recovered haplotypes. The *csp* amplicon worked well at higher parasitemias but tended to overestimate the frequency of the major variant in mixtures. The *trap* amplicon yielded poor results.

Among clinical samples, a total of 4,324,169 reads were extracted across the 4 amplicons, with 2,893,882 reads (66.9%) being used in the final haplotype determination. Thus, the majority of reads recovered were used in the final haplotype determination. This was true on a gene-level basis for three genes, *csp, ama*, and *sera2*. However, for *trap*, the majority of reads failed to produce haplotypes, primarily due to cluster sizes below our cutoff of 500 reads. Details of sequencing yield for each target are shown in **Table 1**. Individual level genotyping data are shown in **Supplemental Table 2**. Unique haplotypes for each gene and the number of samples with those haplotypes are shown in **Supplemental Tables 3-6**.

**Table 1.** Sequencing Success and Characteristics of Targeted Genes.

| Gene | Total Extracted Reads | # Read Used for Final Haplotypes (% of extracted) | Range of Reads per Successful Sample | # of Successful Genotyped Samples | # Haplotypes | He | Tajima's D | Fu and Li D | Fu and Li F | $\pi$ |
| --- | --- | --- | --- | --- | --- | --- | --- | --- | --- | --- |
| *csp* | 192,801 | 140,269 (72.8%) | 516-27,370 | 35 | 22 | 0.908 | 0.22 ($p>0.10$) | 0.24 | 0.23 | 0.021 |
| *ama* | 2,836,417 | 1,837,489 (64.8%) | 554-49,957 | 83 | 35 | 0.952 | 1.94<br>($0.1>p>0.05$) | 1.74 | 2.51 | 0.041 |
| *sera2* | 1,252,534 | 895,025 (71.5%) | 513-49,419 | 75 | 29 | 0.919 | -0.449<br>($p>0.1$) | -0.85 | -0.32 | 0.010 |
| *trap* | 42,417 | 21,099 (49.7%) | 540-7,626 | 11 | 10 | 0.876 | 0.041 ($p>0.1$) | -0.15 | -0.10 | 0.013 |

### Gene Diversity

For *csp*, we successfully genotyped 35/100 (35%) samples (**Table 1**). Twenty-two haplotypes were detected with a heterozygosity of 0.908 in the population (**Supplementary Table 7**). The nucleotide diversity (π) was 0.021. The vaccine haplotype of *csp*, which is identical to the 3D7 strain, is represented by PfCSP.01 in our population. This haplotype was found in 7/35 (20%) of genotyped samples and represented 25,173/140,269 (17.9%) of the sequencing reads used to construct haplotypes.

Genetic diversity was higher, and more samples were successfully genotyped for *ama* and *sera2* (**Table 1**). Thirty-five and 29 haplotypes were detected for *ama* and *sera2*, respectively. Both had higher heterozygosity than *csp*, but π was higher for *ama* and lower for *sera2*. The *trap* locus was the least diverse, with a π of 0.013, 10 haplotypes detected, and a heterozygosity of 0.876. However, it was also the locus with the fewest genotyped samples, with 11 samples genotyped.

### Complexity of Infection

The complexity of infection (COI) was assessed separately for each gene based on the number of haplotypes detected. Among the four genes analyzed, *ama* exhibited the widest range and highest number of haplotypes across samples (**Figure 1**). The proportion of polyclonal infections was 28.6% (10/35) for *csp*, 68.7% (57/83) for *ama*, 65.3% (49/75) for *sera2*, and 18.2% (2/11) for *trap*. Mean COI values (range) were: *csp* – 1.3 (1-2), *ama* – 2.8 (1-9), *sera2* – 2.1 (1-6), and *trap* – 1.2 (1-2). Median COI values (95% CI of the median) were: *csp* – 1 (1–1), *ama* – 2 (2–3), *sera2* – 2 (2–2), and *trap* – 1 (1–2).

**Figure 1.**
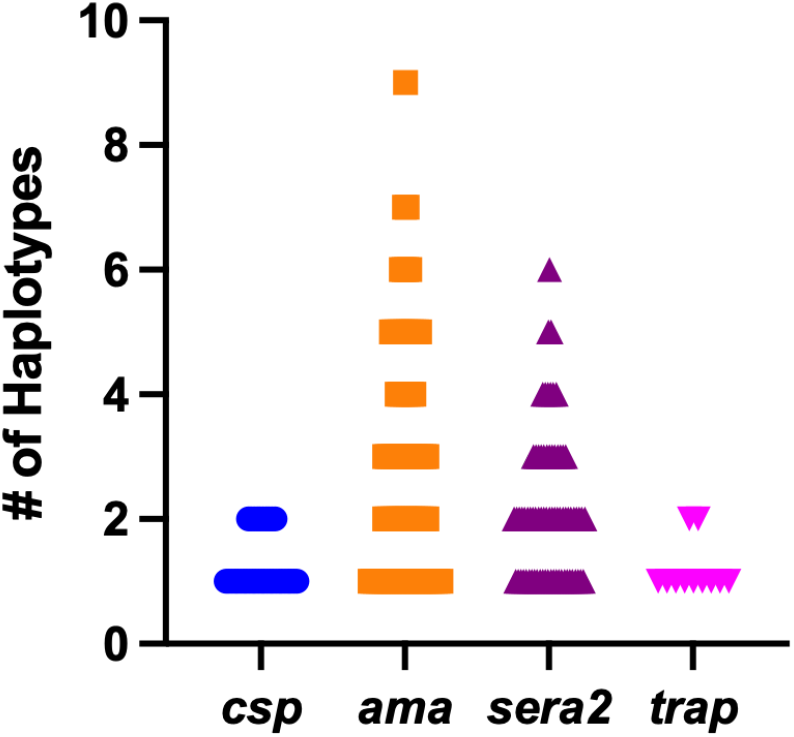
Number of Haplotypes Detected Per Sample at Each Gene. The number of haplotypes detected in individual samples is plotted for *csp* (n=35), *ama* (n=83), *sera2* (n=75), and *trap* (n=11).

### Selection on *csp*

None of the 4 genes showed significant Tajima’s D, Fu and Li D*, or Fu and Li F* values for the complete gene (**Table 1**). The TH2 epitope (amino acids 311–327) and the TH3 epitope (amino acids 352–363) of *csp* showed elevated Tajima’s D values on sliding window analysis (**Figure 2, Panel A**). The translated protein shows high levels of diversity in these regions (**Figure 2, Panel D; Supplemental Figure 1**). Sliding window Tajima’s D analysis shows significantly elevated Tajima’s D at multiple windows in ama, but other genes lack significant windows in either direction (**Supplemental Figure 2**).

**Figure 2.**
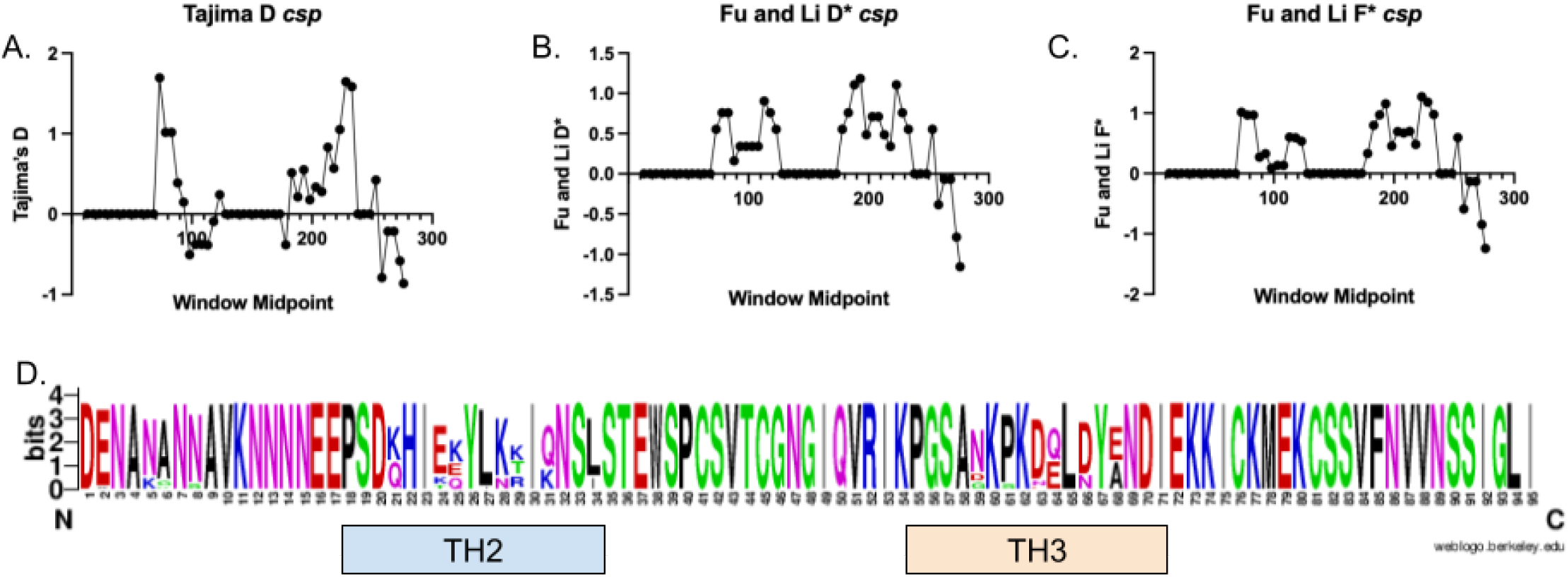
TH2 and TH3 Diversity and Selection in *csp*. Panels A-C show scans for selection across the amplicon using a 25bp widow with a slide of 5bp. No values for Tajima’s D (Panel A), Fu and Li D* (Panel B), or Fu and Li F* (Panel C) reached statistical significance. Panel D shows a web logo of amino acids for the region amplified with the TH2 (amino acids 311–327) region and TH3 region (amino acids 352–363) marked.

### *Diversity of csp* in Cameroon

In order to assess the relationship of *csp* antigen diversity in Kaele to previously published data in Cameroon, we downloaded a subset of sequences from 22 isolates from Pette, also in Northern Cameroon, from a previous report (**Supplemental Table 8**).(Kojom Foko et al., 2024) Data from Efeti et al. was not available for download.(Efeti et al., 2025) Using these data, we generated a median-joining phylogenetic tree of sequences trimmed to the same length (**Figure 3**). The isolates appear to be from a single population, with isolates from both studies intermixed. Identical samples with no branch length differences were found between the studies.

**Figure 3.**
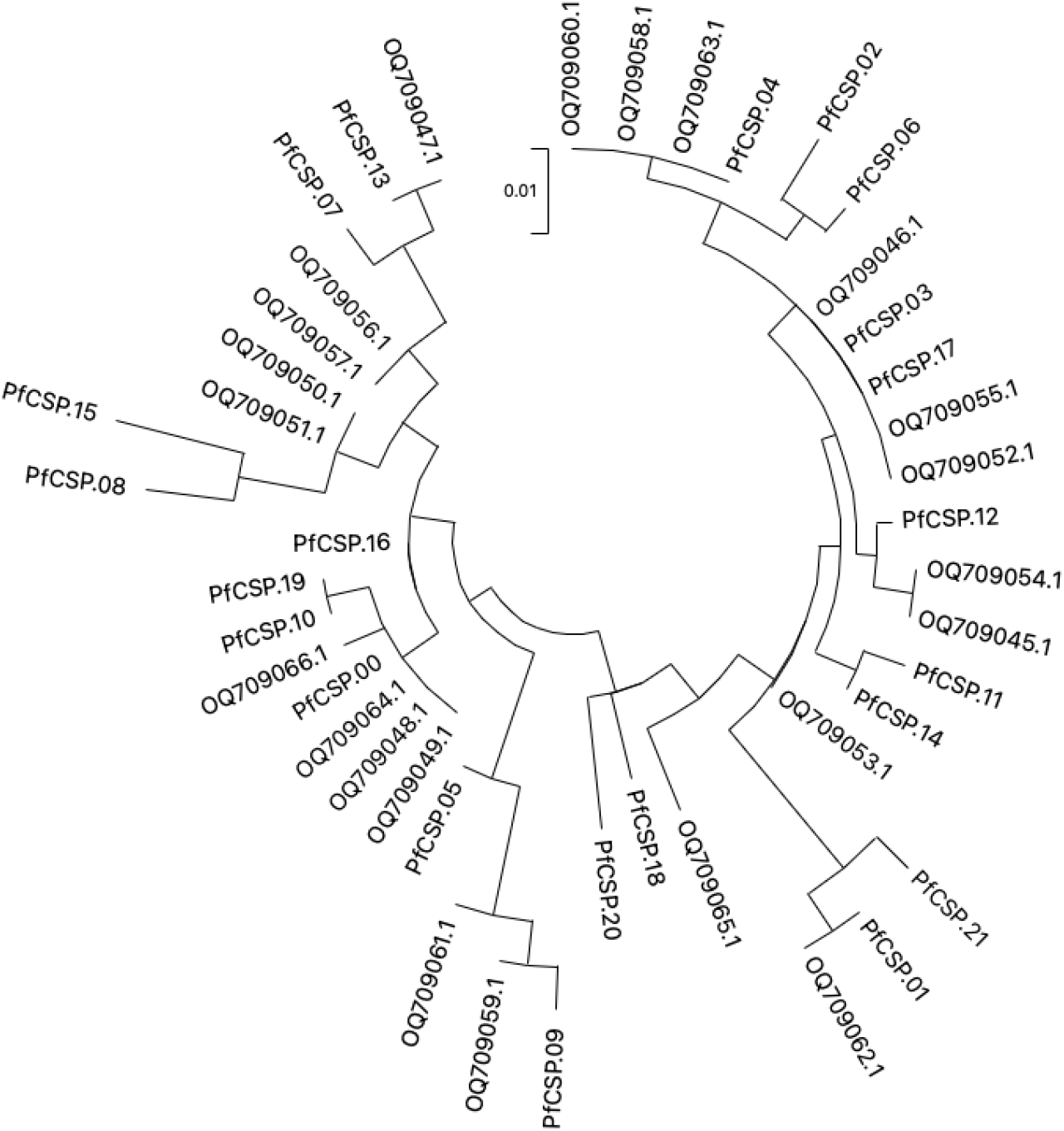
Median Joining Phylogenetic Tree of *csp* Sequences. Twenty-two public sequences from Pette collected in 2019 (start with OQ) were compared to the 22 haplotypes detected in 2022-23 (start with PfCSP)

## DISCUSSION

*P. falciparum* remains a major public health challenge in Cameroon. Despite the availability of effective interventions, the number of malaria cases has been increasing over the last 5 years.(World Health Organization, 2024) Implementation of the malaria vaccine RTS,S/ASO1 has shown promising reductions in the districts where it has been implemented so far, with plans to expand to the entire country.(ReliefWeb, n.d.) Here we describe the diversity of *csp*, the antigen in RTS,S/ASO1, as well as three other malaria antigens (*ama, sera2*, and *trap*), in a northern district of Cameroon prior to vaccine implementation. Given that there is evidence of strain-specific vaccine efficacy, understanding the longitudinal dynamics of the *csp* antigen in the population may be helpful in understanding long-term vaccine efficacy.(Neafsey et al., 2015) We found that the vaccine-matched *csp* haplotype was found in 20% of genotyped infections. The overall diversity of the antigen was similar to previous reports of *csp* diversity in Cameroon.(Kojom Foko et al., 2024) Haplotypes detected in this study were intermixed with haplotypes from Pette in Northern Cameroon, with identical haplotypes detected (**Figure 3**). The TH2 and TH3 epitopes have high levels of diversity and signatures of balancing selection similar to other studies (**Figure 2**).(Aragam et al., 2013)

Consistent with previous studies in Cameroon, we found no strong evidence of evolutionary pressure acting on the C-terminal of the *csp* gene, as Tajima’s D, Fu & Li’s F, and Fu & Li’s D values were not statistically significant.(Kojom Foko et al., 2024) At the level of the complete gene, our results aligned with earlier reports from other countries in Africa as well.(Bailey et al., 2012; Aragam et al., 2013) However, similar to those studies, we observed peaks in Fu & Li’s D*, Fu & Li’s F*, and Tajima’s D around the TH2 and TH3 regions, indicating an excess of intermediate-frequency alleles (**Figure 2**).(Weedall et al., 2007; Bailey et al., 2012; Aragam et al., 2013). These patterns are similar to previously reported work in other regions of Africa.(Bailey et al., 2012; Aragam et al., 2013) This suggests that genetic diversity at these loci is maintained within the population, potentially benefiting the parasite and reflecting weak selective pressure; likely driven by host immunity, as supported by structural modeling.(Bailey et al., 2012; Aragam et al., 2013) Notably, we detected a high proportion (20%) of vaccine-matched infections, significantly exceeding previous reports from Cameroon (2%) and other African sites.(Aragam et al., 2013; Neafsey et al., 2015; Kojom Foko et al., 2024) This discrepancy may be partly due to differences in sequencing methods between the previous study (sanger sequencing) in Cameroon and the present one. Importantly, our sequenced fragment includes the full TH2 and TH3 epitopes, which are known to be under allele-specific selection.

The *csp* C-terminal sequences from our study in Northern Cameroon show strong phylogenetic continuity with those previously reported from the same region (Pette), with isolates from both datasets intermixed and some sequences being identical; This pattern indicates the presence of a single circulating *Plasmodium falciparum* population. Such genetic homogeneity contrasts with the higher diversity observed in southern Cameroonian sites such as Douala, where novel mutations like E357L and D359Y, which may be detrimental based on protein structure, were detected in previous studies.(Kojom Foko et al., 2024) The absence of these mutations in our dataset, along with the low prevalence (<5%) of K322I, suggests limited selective pressure acting on these loci within the northern eco-epidemiological zone (**Supplemental Figure 1**). Importantly, the Cameroonian *csp* sequences have previously been shown to cluster phylogenetically with isolates from Sierra Leone, Ghana, Zambia, and Equatorial Guinea, highlighting regional genetic connectivity within sub-Saharan Africa.(Kojom Foko et al., 2024) Given the close relatedness of our isolates to those from Pette, it is likely that this broader phylogeographic relationship remains consistent.

Other genes showed weak signals of selection, with *ama* being the only locus showing statistically significant selection in sliding window analysis. The positive Tajima’s D values for *ama* are consistent with balancing selection, as reported in other studies.(Osier et al., 2010; Arnott et al., 2014; Naung et al., 2022) This gene also showed the highest nucleotide diversity and second highest number of haplotypes (**Table 1**). This is similar to other reports of *ama* diversity in Cameroon, which reported a similar He (0.976) and a lower nucleotide diversity (0.016)(Hawadak et al., 2024). Tajima’s D in that study was 2.058. Whole genome sequencing work from Mt. Cameroon area showed similar modest levels of balancing selection in *ama* and *trap* (Tajima’s D >1) as shown here.(Apinjoh et al., 2023)

COI estimates from the 4 genes varied, with *ama* providing the highest mean COI estimate, the highest median estimate, and the greatest range of haplotypes detected. Polyclonal infections were commonly detected by *ama* and *sera2*, but not *csp* and *trap*. COI estimates were similar to those estimated by amplicon deep sequencing and molecular inversion probes in Dschang, Cameroon, which estimated COI to be 2.7 (Range 1-5) and 1.2 (range 1-3), respectively.(Sadler et al., 2024) In that study, however, *ama* was not the most informative locus for within sample diversity, providing the most number of haplotypes in 43 of the 100 samples genotyped. Rather, an amplicon at a different hypervariable region (Heome A) provided the highest COI estimate for more samples.(Sadler et al., 2024) The majority of other work in Cameroon regarding COI has relied on the detection of size polymorphisms in merozoite surface protein genes or microsatellites.(Atuh et al., 2022; Biabi et al., 2023; Bouopda-Tuedom et al., 2025) While COI is generally found to be lower with those methods than amplicon deep sequencing, polyclonal infections were commonly found.

Unfortunately, we were only able to generate *csp* genotypes from 35 of 100 samples sequenced, limiting the generalizability of the data and potentially the within-population frequency estimates. Diverse *ama* and *sera2* genes were genotyped from more samples and likely represent the population diversity more accurately. The diversity of *trap* is likely under-estimated due to the low number of samples that were successfully sequenced. We used highly restrictive filtering for determining haplotypes with a high within-sample minimum frequency (>5%) and a high threshold for the number of reads needed (500 reads) to determine a haplotype. This likely impacted both the ability to detect minority variants in mixed infections, as well as the overall success of genotyping samples. Still, this report provides new, important information about parasite antigenic diversity in Cameroon, which can inform vaccine rollout evaluation programs.

In summary, this study provides a critical baseline assessment of *P. falciparum* antigenic diversity and complexity of infection (COI) in Kaelé health district, Cameroon, prior to the implementation of the RTS,S/AS01 malaria vaccine. Our findings reveal substantial diversity in the *csp* gene, particularly within the TH2 and TH3 epitopes, consistent with balancing selection and previous reports from other regions of Africa. Interestingly, 20% of genotyped infections contain a strain that matched the vaccine strain. The overall diversity and phylogenetic relationships suggest a largely homogeneous parasite population structure in Far North Cameroon. The COI estimates underscore the prevalence of polyclonal infections, which may influence vaccine efficacy and parasite evolution. Limitations in genotyping success, particularly for *csp* and *trap*, highlight the need for optimized sequencing approaches to capture minority variants and improve resolution. Despite these constraints, our data contribute valuable insights into the genetic landscape of *P. falciparum* in a vaccine-targeted region and underscore the importance of continued molecular surveillance to monitor antigenic shifts and inform long-term vaccine strategies.

## Acknowledgements

The following reagent was obtained through BEI Resources, NIAID, NIH: *Plasmodium falciparum*, Strain 3D7, MRA-102, contributed by Daniel J. Carucci. The following reagent was obtained through BEI Resources, NIAID, NIH: Genomic DNA from *Plasmodium falciparum*, Strain 3D7, MRA-102G, contributed by Daniel J. Carucci. The following reagent was obtained through BEI Resources, NIAID, NIH: Genomic DNA from *Plasmodium falciparum*, Strain 7G8, MRA-152G, contributed by David Walliker.

## Author Contributions

Data Curation: BV, CCG, JMS, VPKT

Formal Analysis: IMA, JJJ

Funding Acquisition: JJJ, RRD, IMA, SEN

Investigation: BVM, VPKT, CCG, JAS, VVJ, AJM, GYW

Methodology: BVM, JMS, JJJ

Project Administration: IMA, JTL, JJJ

Resources: JJJ, RRD, IMA, SEN, VVJ, AJM, GYW

Software: JAB

Supervision: JJJ, SEN, RRD, JTL, IMA

Validation: JJJ

Visualization: JJJ

Writing – Original Draft Preparation: IMA, VPKT, JJJ

Writing – Review & Editing: All authors

## Data Availability

All data are available within the manuscript. Raw sequencing reads are available in the NCBI SRA (pending).

