## Supplementary material for "Complexity of Infection and *Plasmodium falciparum* Circumsporozoite Protein Diversity Prior to Malaria Vaccine Implementation in Kaelé Health District, Cameroon, 2022-2023": Supplmental Material

### Supplemental Table 1. Control Sample Sequencing

| Control | # reps | # with detected haplotype | Median # of detected haplotypes | Range | # Reps with Haplotype missed | Haplotype 1* | | Haplotype 2* | |
|---|---|---|---|---|---|---|---|---|---|
| | | | | | | Mean Freq. | Range | Mean Freq. | Range |
| **csp** | | | | | | | | | |
| 10 p/µl | 12 | 4 | 1 | 1-1 | 0 | 100% | 100%-100% | - | - |
| 100 p/µl | 12 | 12 | 1 | 1-1 | 0 | 100% | 100%-100% | - | - |
| 1,000 p/µl | 12 | 12 | 1 | 1-1 | 0 | 100% | 100%-100% | - | - |
| Mix 1 (80:20) | 10 | 10 | 2 | 1-2 | 2 | 92.3% | 86.6%-100% | 10.9% | 7.4%-13.4% |
| Mix 2 (95:5) | 10 | 10 | 2 | 1-2 | 5 | 97.5% | 94.4%-100% | 5.0% | 4.5%-5.6% |
| **ama** | | | | | | | | | |
| 10 p/µl | 12 | 7 | 1 | 1-1 | 0 | 100% | 100%-100% | - | - |
| 100 p/µl | 12 | 12 | 1 | 1-1 | 0 | 100% | 100%-100% | - | - |
| 1,000 p/µl | 12 | 12 | 1 | 1-1 | 0 | 100% | 100%-100% | - | - |
| Mix 1 (80:20) | 10 | 10 | 2 | 2-2 | 0 | 77.6% | 73.5%-82.5% | 22.7% | 17.5%-26.5% |
| Mix 2 (95:5) | 10 | 10 | 2 | 1-2 | 2 | 97.0% | 95.3%-98.7% | 3.8% | 2.6%-4.9% |
| **sera2** | | | | | | | | | |
| 10 p/µl | 12 | 6 | 1 | 1-1 | 0 | 100% | 100%-100% | - | - |
| 100 p/µl | 12 | 12 | 1 | 1-1 | 0 | 100% | 100%-100% | - | - |
| 1,000 p/µl | 12 | 12 | 1 | 1-1 | 0 | 100% | 100%-100% | - | - |
| Mix 1 (80:20) | 10 | 10 | 2 | 2-2 | 0 | 79.2% | 76.7%-81.5% | 20.8% | 18.5%-23.9% |
| Mix 2 (95:5) | 10 | 10 | 2 | 2-2 | 0 | 93.5% | 93.1%-94.7% | 6.5% | 5.3%-6.9% |
| **trap** | | | | | | | | | |
| 10 p/µl | 12 | 0 | - | - | - | - | - | - | - |
| 100 p/µl | 12 | 12 | 1 | 1-1 | 0 | 100% | 100%-100% | - | - |
| 1,000 p/µl | 12 | 12 | 1 | 1-1 | 0 | 100% | 100%-100% | - | - |
| Mix 1 (80:20) | 10 | 9 | 2 | 1-3 | 1[#] | 82.7% | 76.3%-83.0% | 16.8% | 12.4%-19.3% |
| Mix 2 (95:5) | 10 | 10 | 1 | 1-1 | 10 | 100% | 100%-100% | - | - |

*: Of replicates with detectable haplotypes (zeros not counted).

#: Two samples with a false minor variant. Both were the same haplotype at 10.8% and 10.3% within control frequency. The haplotype contained 4 mutations, the first 2 were 3D7-like, the second 2 were 7G8-like, suggesting this may be a chimera. This haplotype was not found in any samples.

**Supplemental Table 2. Individual Level Genotyping Data.**

Please see uploaded Excel sheet.

**Supplemental Table 3. *csp* Haplotypes Detected**

| Haplotype | # Samples Detected | Sequence |
|---|---|---|
| PfCSP.00 | 9 | TATTAATCCTATTGAACTATTTACGACATTAAACACACTGGAACATTTTTCCATTTTACAAATTTTTTTTTCAATATCATTTTCATAATCTAATTGGTCTTTAGG<br>TTTATTAGCAGAGCCAGGCTTTATTCTAACTTGAATACCATTTCCACAAGTTACACTACATGGGGACCATTCAGTTGAAAGAGAATTTTGTATTGTCTTTAA<br>ATATTGTTCTATGTGCTTATCACTTGGTTCTTCGTTATTATTATTTTTTACAGCATTGTTGGCTTTAGCATTTTCATCT |
| PfCSP.01 | 7 | TATTAATCCTATTGAACTATTTACGACATTAAACACACTGGAACATTTTTCCATTTTACAAATTTTTTTTTCAATATCATTTGCATAATCTAATTCGTCTTTAGG<br>TTTATTAGCAGAGCCAGGCTTTATTCTAACTTGAATACCATTTCCACAAGTTACACTACATGGGGACCATTCAGTTGAAAGAGAATTTTGTATTTTGTTTAA<br>ATATTCTTTTATGTGCTTATCACTTGGTTCTTCGTTATTATTATTTTTTACAGCACTGTTGGCATTAGCATTTTCATCT |
| PfCSP.02 | 4 | TATTAATCCTATTGAACTATTTACGACATTAAACACACTGGAACATTTTTCCATTTTACAAATTTTTTTTTCAATATCATTTGCATAATCTAATTGGTCTTTAGG<br>TTTATTAGCAGAGCCAGGCTTTATTCTAACTTGAATACCATTTCCACAAGTTACACTACATGGGGACCATTCAGTTGAAAGAGAATTTTTTATTTTGTTTAA<br>ATATTTTTCTATGTGCTGATCACTTGGTTCTTCGTTATTATTATTTTTTACAGCATTGTTGGCATTAGCATTTTCATCT |
| PfCSP.03 | 3 | TATTAATCCTATTGAACTATTTACGACATTAAACACACTGGAACATTTTTCCATTTTACAAATTTTTTTTTCAATATCATTTGCATAATCTAATTGGTCTTTAGG<br>TTTATTAGCAGAGCCAGGCTTTATTCTAACTTGAATACCATTTCCACAAGTTACACTACATGGGGACCATTCAGTTGAAAGAGAATTTTGTATTGTCTTTAA<br>ATATTTTTCTATGTGCTGATCACTTGGTTCTTCGTTATTATTATTTTTTACAGCATTGTTGGCATTAGCATTTTCATCT |
| PfCSP.04 | 3 | TATTAATCCTATTGAACTATTTACGACATTAAACACACTGGAACATTTTTCCATTTTACAAATTTTTTTTTCAATATCATTTGCATAATCTAATTGGTCTTTAGG<br>TTTATTAGCAGAGCCAGGCTTTATTCTAACTTGAATACCATTTCCACAAGTTACACTACATGGGGACCATTCAGTTGAAAGAGAATTTTTTATTGTCTTTAA<br>ATATTTTTCTATGTGCTGATCACTTGGTTCTTCGTTATTATTATTTTTTACAGCATTGTTGGCATTAGCATTTTCATCT |
| PfCSP.05 | 2 | TATTAATCCTATTGAACTATTTACGACATTAAACACACTGGAACATTTTTCCATTTTACAAATTTTTTTTTCAATATCATTTTCATAATCTAATTCGTCTTTAGA<br>TTTACCAGCAGAGCCAGGCTTTATTCTAACTTGAATACCATTTCCACAAGTTACACTACATGGGGACCATTCAGTTGAAAGAGAATTTTGTATTTTCTTTA<br>AATATTGTTCTATGTGCTTATCACTTGGTTCTTCGTTATTATTATTTTTTACAGCATTGTTGGCATTAGCATTTTCATCT |
| PfCSP.06 | 2 | TATTAATCCTATTGAACTATTTACGACATTAAACACACTGGAACATTTTTCCATTTTACAAATTTTTTTTTCAATATCATTTGCATAATCTAATTGGTCTTTAGG<br>TTTATTAGCAGAGCCAGGCTTTATTCTAACTTGAATACCATTTCCACAAGTTACACTACATGGGGACCATTCAGTTGAAAGAGAATTTTTTATTATCTTTAA<br>ATATTTTTCTATGTGCTGATCACTTGGTTCTTCGTTATTATTATTTTTTACAGCATTGTTGGCATTAGCATTTTCATCT |
| PfCSP.07 | 1 | TATTAATCCTATTGAACTATTTACGACATTAAACACACTGGAACATTTTTCCATTTTACAAATTTTTTTTTCAATATCATTTTCATAATTTAATTCGTCTTTAGG<br>TTTATTAGCAGAGCCAGGCTTTATTCTAACTTGAATACCATTTCCACAAGTTACACTACATGGGGACCATTCAGTTGAAATAGAATTTTTTATTTTGTTTAA<br>ATATTGTTCTATGTGCTTATCACTTGGTTCTTCGTTATTATTATTTTTTACAGCATTGTTGGCATTAGCATTTTCATCT |
| PfCSP.08 | 1 | TATTAATCCTATTGAACTATTTACGACATTAAACACACTGGAACATTTTTCCATTTTACAAATTTTTTTTTCAATATCATTTTCATAATTTAATTCCTCTTTAGG<br>TTTATTAGCAGAGCCAGGCTTTATTCTAACTTGAATACCATTTCCACAAGTTACACTACATGGGGACCATTCAGTTGAAAGAGAATTTTGTATTCTCTTTAA<br>ATATTCTGTTATGTGCTTATCACTTGGTTCTTCGTTATTATTATTTTTTACAGCATTGTTGGAATTAGCATTTTCATCT |
| PfCSP.09 | 1 | TATTAATCCTATTGAACTATTTACGACATTAAACACACTGGAACATTTTTCCATTTTACAAATTTTTTTTTCAATATCATTTTCATAATCTAATTCGTTTTTAGAT<br>TTACCAGCAGAGCCAGGCTTTATTCTAACTTGAATACCATTTCCACAAGTTACACTACATGGGGACCATTCAGTTGAAAGAGAATTTTGTATTCTCTTTAA<br>ATATTCTGTTATGTGCTTATCACTTGGTTCTTCGTTATTATTATTTTTTACAGCATTGTTGCCATTAGCATTTTCATCT |
| PfCSP.10 | 1 | TATTAATCCTATTGAACTATTTACGACATTAAACACACTGGAACATTTTTCCATTTTACAAATTTTTTTTTCAATATCATTTTCATAATCTAATTGGTCTTTAGG<br>TTTATTAGCAGAGCCAGGCTTTATTCTAACTTGAATACCATTTCCACAAGTTACACTACATGGGGACCATTCAGTTGAAAGAGAATTTTTTATTGTCTTTAA |

| | | |
|---|---|---|
| | | ATATTGTTCTATGTGCTTATCACTTGGTTCTTCGTTATTATTATTTTTTACAGCATTGTTGGCTTTAGCATTTTCATCT |
| PfCSP.11 | 1 | TATTAATCCTATTGAACTATTTACGACATTAAACACACTGGAACATTTTTCCATTTTACAAATTTTTTTTTCAATATCATTTGCATAATCTAATTGGTCTTTAGG<br>TTTATCAGCAGAGCCAGGCTTTATTCTAACTTGAATACCATTTCCACAAGTTACACTACATGGGGACCATTCAGTTGAAAGAGAATTTTGTATTCTCTTTA<br>AATATTTTTCTATGTGCTGATCACTTGGTTCTTCGTTATTATTATTTTTTACAGCATTGTTGGCATTAGCATTTTCATCT |
| PfCSP.12 | 1 | TATTAATCCTATTGAACTATTTACGACATTAAACACACTGGAACATTTTTCCATTTTACAAATTTTTTTTTCAATATCATTTGCATAATCTAATTGGTCTTTAGG<br>TTTATTAGCAGAGCCAGGCTTTATTCTAACTTGAATACCATTTCCACAAGTTACACTACATGGGGACCATTCAGTTGAAAGAGAATTTTGTATTCTCTTTAA<br>ATATTTTTCTATGTGCTGATCACTTGGTTCTTCGTTATTATTATTTTTTACAGCATTGTTGGCATTAGCATTTTCATCT |
| PfCSP.13 | 1 | TATTAATCCTATTGAACTATTTACGACATTAAACACACTGGAACATTTTTCCATTTTACAAATTTTTTTTTCAATATCATTTTCATAATTTAATTCGTCTTTAGG<br>TTTATTAGCAGAGCCAGGCTTTATTCTAACTTGAATACCATTTCCACAAGTTACACTACATGGGGACCATTCAGTTGAAATAGAATTTTTTATTTTCTTTAAA<br>TATTCTTCTATGTGCTTATCACTTGGTTCTTCGTTATTATTATTTTTTACAGCATTGTTGGCATTAGCATTTTCATCT |
| PfCSP.14 | 1 | TATTAATCCTATTGAACTATTTACGACATTAAACACACTGGAACATTTTTCCATTTTACAAATTTTTTTTTCAATATCATTTGCATAATCTAATTGGTCTTTAGG<br>TTTATCAGCAGAGCCAGGCTTTATTCTAACTTGAATACCATTTCCACAAGTTACACTACATGGGGACCATTCAGTTGAAAGAGAATTTTGTATTTTCTTTAA<br>ATATTTTTCTATGTGCTGATCACTTGGTTCTTCGTTATTATTATTTTTTACAGCATTGTTGCCATTAGCATTTTCATCT |
| PfCSP.15 | 1 | TATTAATCCTATTGAACTATTTACGACATTAAACACACTGGAACATTTTTCCATTTTACAAATTTTTTTTTCAATATCATTTTCATAATTTAATTCGTCTTTAGG<br>TTTACCAGCAGAGCCAGGCTTTATTCTAACTTGAATACCATTTCCACAAGTTACACTACATGGGGACCATTCAGTTGAAATAGAATTTTGTATTCTCTTTAA<br>ATATTCTTTTATGTGCTTATCACTTGGTTCTTCGTTATTATTATTTTTTACAGCATTGTTGGCATTAGCATTTTCATCT |
| PfCSP.16 | 1 | TATTAATCCTATTGAACTATTTACGACATTAAACACACTGGAACATTTTTCCATTTTACAAATTTTTTTTTCAATATCATTTTCATAATCTAATTCGTCTTTAGG<br>TTTATTAGCAGAGCCAGGCTTTATTCTAACTTGAATACCATTTCCACAAGTTACACTACATGGGGACCATTCAGTTGAAAGAGAATTTTGTATTGTCTTTAA<br>ATATTGTTCTATGTGCTTATCACTTGGTTCTTCGTTATTATTATTTTTTACAGCATTGTTGGCTTTAGCATTTTCATCT |
| PfCSP.17 | 1 | TATTAATCCTATTGAACTATTTACGACATTAAACACACTGGAACATTTTTCCATTTTACAAATTTTTTTTTCAATATCATTTGCATAATCTAATTGGTCTTTAGG<br>TTTATTAGCAGAGCCAGGCTTTATTCTAACTTGAATACCATTTCCACAAGTTACACTACATGGGGACCATTCAGTTGAAAGAGAATTTTGTATTGTCTTTAA<br>ATATTTTTCTATGTGCTGATCACTTGGTTCTTCGTTATTATTATTTTTTACAGCATTGTTGGCTTTAGCATTTTCATCT |
| PfCSP.18 | 1 | TATTAATCCTATTGAACTATTTACGACATTAAACACACTGGAACATTTTTCCATTTTACAAATTTTTTTTTCAATATCATTTTCATAATTTAATTCGTCTTTAGG<br>TTTATTAGCAGAGCCAGGCTTTATTCTAACTTGAATACCATTTCCACAAGTTACACTACATGGGGACCATTCAGTTGAAAGAGAATTTTGTATTCTCTTTAA<br>ATATTTTTCTATGTGCTGATCACTTGGTTCTTCGTTATTATTATTTTTTACAGCATTGTTGGCATTAGCATTTTCATCT |
| PfCSP.19 | 1 | TATTAATCCTATTGAACTATTTACGACATTAAACACACTGGAACATTTTTCCATTTTACAAATTTTTTTTTCAATATCATTTTCATAATCTAATTGGTCTTTAGG<br>TTTATTAGCAGAGCCAGGCTTTATTCTAACTTGAATACCATTTCCACAAGTTACACTACATGGGGACCATTCAGTTGAAAGAGAATTTTTTATTGTCTTTAA<br>ATATTGTTCTATGTGCTTATCACTTGGTTCTTCGTTATTATTATTTTTTACAGCATTGTTGGCATTAGCATTTTCATCT |
| PfCSP.20 | 1 | TATTAATCCTATTGAACTATTTACGACATTAAACACACTGGAACATTTTTCCATTTTACAAATTTTTTTTTCAATATCATTTTCATAATCTAATTCGTTTTTAGG<br>TTTATCAGCAGAGCCAGGCTTTATTCTAACTTGAATACCATTTCCACAAGTTACACTACATGGGGACCATTCAGTTGAAAGAGAATTTTGTATTTTCTTTAT<br>ATATTTTTCTATGTGCTGATCACTTGGTTCTTCGTTATTATTATTTTTTACAGCATTGTTGGCATTAGCATTTGCATCT |
| PfCSP.21 | 1 | TATTAATCCTATTGAACTATTTACGACATTAAACACACTGGAACATTTTTCCATTTTACAAATTTTTTTTTCAATATCATTTGCATAATCTAATTGGTCTTTAGG<br>TTTATTAGCAGAGCCAGGCTTTATTCTAACTTGAATACCATTTCCACAAGTTACACTACATGGGGACCATTCAGTTGAAATAGAATTTTGTATTTTGTTTAA<br>ATATTCTTTTATGTGCTTATCACTTGGTTCTTCGTTATTATTATTTTTTACAGCACTGTTGGCATTAGCATTTTCATCT |

**Supplemental Table 4. *ama* Haplotypes Detected**

| Haplotype | # Samples Detected | Sequence |
|---|---|---|
| PfAMA1.00 | 23 | TTTGGTAAAGGTATAATTATTGAGAATTCAAATACTACTTTTTTAAAACCGGTAGCTACGGGAAATCAAGATTTAAAAGATGGAGGTTTTGCTTTTCCTCCAACAGAACCTCTTATATCACCAATGACATTAAATGGTATGAGAGATTTTTATAAAAATAATGAATATGTAAAAAATTTAGATGAATTGACTTT |
| PfAMA1.01 | 18 | TTTGGTAAAGGTATAATTATTGAGAATTCAAATACTACTTTTTTAACACCGGTAGCTACGGGAAAACAAGATTTAAAAGATGGAGGTTTTGCTTTTCCTCCAACAAATCCTCTTATATCACCAATGACATTAAATGGTATGAAAGATTTTTATAAAGATAATGAAGATGTAAAAAATTTAGATGAATTGACTTT |
| PfAMA1.02 | 17 | TTTGGTAAAGGTATAATTATTGAGAATTCAAAAACTACTTTTTTAACACCGGTAGCTACGGAAAATCAAGATTTAAAAGATGGAGGTTTTGCTTTTCCTCCAACAAATCCTCCTATGTCACCAATGACATTAAATGGTATGAGAGATTTATATAAAAATAATGAATATGTAAAAAATTTAGATGAATTGACTTT |
| PfAMA1.03 | 16 | TTTGGTAAAGGTATAATTATTGAGAATTCAAATACTACTTTTTTAACACCGGTAGCTACGGGAAATCAAGATTTAAAAGATGGAGGTTTTGCTTTTCCTCCAACAAAACCTCTTATGTCACCAATGACATTAGATGATATGAGACTTTTGTATAAAGATAATGAAGATGTAAAAAATTTAGATGAATTGACTTT |
| PfAMA1.04 | 15 | TTTGGTAAAGGTATAATTATTGAGAATTCAAATACTACTTTTTTAACACCGGTAGCTACGGAAAATCAAGATTTAAAAGATGGAGGTTTTGCTTTTCCTCCAACAAAACCTCTTATGTCACCAATGACATTAGATCAAATGAGACATTTTTATAAAGATAATAAATATGTAAAAAATTTAGATGAATTGACTTT |
| PfAMA1.05 | 12 | TTTGGTAAAGGTATAATTATTGAGAATTCAAATACTACTTTTTTAACACCGGTAGCTACGGGAAATCAATATTTAAAAGATGGAGGTTTTGCTTTTCCTCCAACAGAACCTCTTATGTCACCAATGACATTAGATGAAATGAGACATTTTTATAAAGATAATAAATATGTAAAAAATTTAGATGAATTGACTTT |
| PfAMA1.06 | 11 | TTTGGTAAAGGTATAATTATTGAGAATTCAAATACTACTTTTTTAACACCGGTAGCTACGGAAAATCAAGATTTAAAAGATGGAGGTTTTGCTTTTCCTCCAACAAATCCTCTTATATCACCAATGACATTAGATCATATGAGAGATTCTTATAAAAATAATGAATATGTAAAAAATTTAGATGAATTGACTTT |
| PfAMA1.07 | 11 | TTTGGTAAAGGTATAATTATTGAGAATTCAAATACTACTTTTTTAACACCGGTAGCTACGGAAAATCAAGATTTAAAAGATGGAGGTTTTGCTTTTCCTCCAACAGAACCTCTTATGTCACCAATGACATTAGATCAAATGAGACATTTTTATAAAGATAATAAATATGTAAAAAATTTAGATGAATTGACTTT |
| PfAMA1.08 | 11 | TTTGGTAAAGGTATAATTATTGAGAATTCAAAAACTACTTTTTTAACACCGGTAGCTACGGAAAATCAAGATTTAAAAGATGGAGGTTTTGCTTTTCCTCCAACAGAACCTCTTATGTCACCAATGACATTAGATGATATGAGACGTTTTTATAAAGATAATGAATATGTAAAAAATTTAGATGAATTGACTTT |
| PfAMA1.09 | 10 | TTTGGTAAAGGTATAATTATTGAGAATTCAAATACTACTTTTTTAACACCGGTAGCTACGGAAAATCAAGATTTAAAAGATGGAGGTTTTGCTTTTCCTCCAACAAAACCTCTTATATCACCAATGACATTAGATCAAATGAGAGATTTATATAAAAATAATGAATATGTAAAAAATTTAGATGAATTGACTTT |
| PfAMA1.10 | 9 | TTTGGTAAAGGTATAATTATTGAGAATTCAAATACTACTTTTTTAACACCGGTAGCTACGGAAAATCAAGATTTAAAAGATGGAGGTTTTGCTTTTCCTCCAACAAAACCTCTTATGTCACCAATGACATTAGATCAAATGAGAGATTTTTATAAAAATAATGAATATGTAAAAAATTTAGATGAATTGACTTT |
| PfAMA1.11 | 9 | TTTGGTAAAGGTATAATTATTGAGAATTCAAAAACTACTTTTTTAACACCGGTAGCTACGGAAAATCAAGATTTAAAAGATGGAGGTTTTGCTTTTCCTCCAACAAAACCTCTTATGTCACCAATGACATTAGATGATATGAGACTTTTGTATAAAGATAATGAAGATGTAAAAAATTTAGATGAATTGACTTT |
| PfAMA1.12 | 6 | TTTGGTAAAGGTATAATTATTGAGAATTCAAATACTACTTTTTTAACACCGGTAGCTACGGAAAATCAAGATTTAAAAGATGGAGGTTTTGCTTTTCCTCCAACAAATCCTCCTATGTCACCAATGACATTAGATCAAATGAGACATTTTTATAAAGATAATAAATATGTAAAAAATTTAGATGAATTGACTTT |
| PfAMA1.13 | 6 | TTTGGTAAAGGTATAATTATTGAGAATTCAAATACTACTTTTTTAACACCGGTAGCTACGGAAAATCAAGATTTAAAAGATGGAGGTTTTGCTTTTCCTCCAACAAATCCTCTTATATCACCAATGACATTAAATGGTATGAAAGATTTTTATAAAGATAATGAAGATGTAAAAAATTTAGATGAATTGACTTT |
| PfAMA1.14 | 6 | TTTGGTAAAGGTATAATTATTGAGAATTCAAATACTACTTTTTTAAAACCGGTAGCTACGGGAAATCAAGATTTAAAAGATGGAGGTTTTGCTTTTCCTCCAACAAATCCTCTTATATCACCAATGACATTAGATCAAATGAGACATTTTTATAAAGATAATGAAGATGTAAAAAATTTAGATGAATTGACTTT |

| | | |
|---|---|---|
| PfAMA1.15 | 5 | TTTGGTAAAGGTATAATTATTGAGAATTCAAATACTACTTTTTTAACACCGGTAGCTACGGGAAAACAAGATTTAAAAGATGGAGGTTTTGCTTTTCCTC<br>CAACAAATCCTCTTATATCACCAATGACATTAGATCATATGAGAGATTTTTATAAAAAAAATGAATATGTAAAAAATTTAGATGAATTGACTTT |
| PfAMA1.16 | 5 | TTTGGTAAAGGTATAATTATTGAGAATTCAAATACTACTTTTTTAACACCGGTAGCTACGGAAAATCAAGATTTAAAAGATGGAGGTTTTGCTTTTCCTCC<br>AACAAAACCTCTTATGTCACCAATGACATTAGATGAAATGAGACATTTTTATAAAGATAATAAATATGTAAAAAATTTAGATGAATTGACTTT |
| PfAMA1.17 | 5 | TTTGGTAAAGGTATAATTATTGAGAATTCAAAAACTACTTTTTTAACACCGGTAGCTACGGAAAATCAAGATTTAAAAGATGGAGGTTTTGCTTTTCCTC<br>CAACAAATCCTCCTATGTCACCAATGACATTAGATGATATGAGACTTTTGTATAAAGATAATGAAGATGTAAAAAATTTAGATGAATTGACTTT |
| PfAMA1.18 | 4 | TTTGGTAAAGGTATAATTATTGAGAATTCAAATACTACTTTTTTAAAACCGGTAGCTACGGAAAATCAAGATTTAAAAGATGGAGGTTTTGCTTTTCCTCC<br>AACAAATCCTCTTATGTCACCAATGACATTAGATCATATGAGACATCTTTATAAAGATAATGAATATGTAAAAAATTTAGATGAATTGACTTT |
| PfAMA1.19 | 4 | TTTGGTAAAGGTATAATTATTGAGAATTCAAATACTACTTTTTTAAAACCGGTAGCTACGGGAAATCAAGATTTAAAAGATGGAGGTTTTGCTTTTCCTCC<br>AACAGAACCTCTTATATCACCAATGACATTAGATGATATGAGAGATTTTTATAAAAATAATGAATATGTAAAAAATTTAGATGAATTGACTTT |
| PfAMA1.20 | 4 | TTTGGTAAAGGTATAATTATTGAGAATTCAAATACTACTTTTTTAACACCGGTAGCTACGGAAAATCAAGATTTAAAAGATGGAGGTTTTGCTTTTCCTCC<br>AACAGAACCTCTTATGTCACCAATGACATTAGATCGTATGAGAGATTTTTATAAAAATAATGAAGATGTAAAAAATTTAGATGAATTGACTTT |
| PfAMA1.21 | 4 | TTTGGTAAAGGTATAATTATTGAGAATTCAAATACTACTTTTTTAACACCGGTAGCTACGGGAAATCAATATTTAAAAGATGGAGGTTTTGCTTTTCCTCC<br>AACAGAACCTCATATGTCACCAATGACATTAGATGAAATGAGACATTTTTATAAAGATAATAAATATGTAAAAAATTTAGATGAATTGACTTT |
| PfAMA1.22 | 3 | TTTGGTAAAGGTATAATTATTGAGAATTCAAATACTACTTTTTTAACACCGGTAGCTACGGGAAAACAAGATTTAAAAGATGGAGGTTTTGCTTTTCCTC<br>CAACAAATCCTCTTATATCACCAATGACATTAAATGGTATGAGAGATTTATATAAAAATAATGAAGATGTAAAAAATTTAGATGAATTGACTTT |
| PfAMA1.23 | 3 | TTTGGTAAAGGTATAATTATTGAGAATTCAAATACTACTTTTTTAAAACCGGTAGCTACGGGAAATCAAGATTTAAAAGATGGAGGTTTTGCTTTTCCTCC<br>AACAAATCCTCTTATATCACCAATGACATTAGATCATATGAGAGATTTTTATAAAAATAATGAATATGTAAAAAATTTAGATGAATTGACTTT |
| PfAMA1.24 | 3 | TTTGGTAAAGGTATAATTATTGAGAATTCAAATACTACTTTTTTAAAACCGGTAGCTACGGGAAATCAAGATTTAAAAGATGGAGGTTTTGCTTTTCCTCC<br>AACAGAACCTCTTATATCACCAATGACATTAAAGGGTATGAGAGATTTTTATAAAAATAATGAATATGTAAAAAATTTAGATGAATTGACTTT |
| PfAMA1.25 | 2 | TTTGGTAAAGGTATAATTATTGAGAATTCAAATACTACTTTTTTAACACCGGTAGCTACGGAAAATCAAGATTTAAAAGATGGAGGTTTTGCTTTTCCTCC<br>AACAAAACCTCTTATGTCACCAATGACATTAGATGATATGAGACTTTTGTATAAAGATAATGAAGATGTAAAAAATTTAGATGAATTGACTTT |
| PfAMA1.26 | 2 | TTTGGTAAAGGTATAATTATTGAGAATTCAAATACTACTTTTTTAACACCGGTAGCTACGGAAAATCAAGATTTAAAAGATGGAGGTTTTGCTTTTCCTCC<br>AACAAAACCTCATATGTCACCAATGACATTAGATGATATGAGACTTTTGTATAAAGATAATGAAGATGTAAAAAATTTAGATGAATTGACTTT |
| PfAMA1.27 | 2 | TTTGGTAAAGGTATAATTATTGAGAATTCAAATACTACTTTTTTAAAACCGGTAGCTACGGGAAATCAAGATTTAAAAGATGGAGGTTTTGCTTTTCCTCC<br>AACAAATCCTCTTATATCACCAATGACATTAAATGGTATGAAAGATTTTTATAAAGATAATGAAGATGTAAAAAATTTAGATGAATTGACTTT |
| PfAMA1.28 | 2 | TTTGGTAAAGGTATAATTATTGAGAATTCAAAAACTACTTTTTTAACACCGGTAGCTACGGAAAATCAAGATTTAAAAGATGGAGGTTTTGCTTTTCCTC<br>CAACAAAACCTCTTATGTCACCAATGACATTAGATGATATGAGAGATTTTTATAAAGATAATGAATATGTAAAAAATTTAGATGAATTGACTTT |
| PfAMA1.29 | 2 | TTTGGTAAAGGTATAATTATTGAGAATTCAAATACTACTTTTTTAAAACCGGTAGCTACGGGAAATCAAGATTTAAAAGATGGAGGTTTTGCTTTTCCTCC<br>AACAAATCCTCTTATATCACCAATGACATTAAATGGTATGAGAGATTTATATAAAAATAATGAAGATGTAAAAAATTTAGATGAATTGACTTT |
| PfAMA1.30 | 2 | TTTGGTAAAGGTATAATTATTGAGAATTCAAAAACTACTTTTTTAACACCGGTAGCTACGGAAAATCAAGATTTAAAAGATGGAGGTTTTGCTTTTCCTC<br>CAACAGAACCTCCTATGTCACCAATGACATTAGATGAAATGAGACATTTTTATAAAGATAATAAATATGTAAAAAATTTAGATGAATTGACTTT |
| PfAMA1.31 | 1 | TTTGGTAAAGGTATAATTATTGAGAATTCAAATACTACTTTTTTAAAACCGGTAGCTACGGGAAATCAAGATTTAAAAGATGGAGGTTTTGCTTTTCCTCC<br>AACAAATCCTCTTATATCACCAATGACATTAGATCATATGAGAGATTTTTATAAAAAAAATGAATATGTAAAAAATTTAGATGAATTGACTTT |

| | | |
|---|---|---|
| PfAMA1.32 | 1 | TTTGGTAAAGGTATAATTATTGAGAATTCAAATACTACTTTTTTAAAACCGGTAGCTACGGAAAATCAAGATTTAAAAGATGGAGGTTTTGCTTTTCCTCC<br>AACAGAACCTCTTATATCACCAATGACATTAGATGATATGAGAGATTTTTATAAAAATAATGAATATGTAAAAAATTTAGATGAATTGACTTT |
| PfAMA1.33 | 1 | TTTGGTAAAGGTATAATTATTGAGAATTCAAATACTACTTTTTTAAAACCGGTAGCTACGGGAAATCAACATTTAAAAGATGGAGGTTTTGCTTTTCCTCC<br>AACAGAACCTCTTATATCACCAATGACATTAGATGATATGAGAGATTTTTATAAAAATAATGAATATGTAAAAAATTTAGATGAATTGACTTT |
| PfAMA1.34 | 1 | TTTGGTAAAGGTATAATTATTGAGAATTCAAAAACTACTTTTTTAACACCGGTAGCTACGGAAAATCAAGATTTAAAAGATGGAGGTTTTGCTTTTCCTC<br>CAACAAAACCTCTTATGTCACCAATGACATTAGATGATATGAGACTTTTGTATAAAGATAATGAATATGTAAAAAATTTAGATGAATTGACTTT |

**Supplemental Table 5. *sera2* Haplotypes Detected**

| Haplotype | # Samples Detected | Sequence |
|---|---|---|
| PfSERA2.00 | 29 | TCCAGGTGATATCTTTGTTCCTACGGATCCTCCTGTTCCTGATGATCTTCCTGCTCTTTCGACATCTGATTGGGATACTTCTACACCTGATTTTGCTGATTC<br>TACTTTTTGTTCTGCTCCTATACCACTTCCTACTTCTTTTTGTTGTTGCTGTTGTTGGGGTTGTGTTTCTTGAGCTAAAGTTGGTAATGCTGGTTGTTGTTTT<br>GGTTTTTCAATTCTTGCTTGTGTTGTTACACGTGAACCATCGGATGATAATG |
| PfSERA2.01 | 22 | TCCAGGTGATATCTTTGTTCCTACGGATCCTCCTGTTCCTGATGATCTTCCTGCTCTTTCGACATCTGATTGGGATACTTCTACACCTGGTTTTGCTGATTC<br>TACTTTTTGTTCTGCTCCTATACCACTTCCTACTTCTTTTTGTTGTTGCTGTTGTTGGGGTTGTGTTTCTTGAGCTAAAGTTGGTAATGCTGGTTGTTGTTTT<br>GGTTTTTCAATTCTTGCTTGTGTTGTTACACGTGAACCATCGGATGATAATG |
| PfSERA2.02 | 13 | TCCAGGTGATATCTTTGTTCCTACGGATCCTCCTGTTCCTGATGATCTTCCTGCTCTTTCGACATCTGATTGGGATACTTCTGCACCTGGTTTTGCTGATTC<br>TACTTTTTGTTCTGCTCCTATACCACTTCCTACTTCTTTTTGTTGTTGCTGTTGTTGGGGTTGTGTTTCTTGAGCTAAAGTTGGTAATGCTGGTTGTTGTTTT<br>GGTTTTTCAATTCTTGCTTGTGTTGTTACACGTGAACCATCGGATGATAATG |
| PfSERA2.03 | 12 | TCCAGGTGATATCTTTGTTCCTACGGATCCTCCTGTTCCTGATGATCTTCCTGCTCTTTCGACATCTGATTGGGATACTTCTGCACCTGGTCTTGCTGATTC<br>TACTTTTTGTTCTGCTCCTATACCACTTCCTACTTCTTTTTGTTGTTGCTGTTGTTGGGGTTGTGTTTCTTGAGCTAAAGTTGGTAATGTTGGTTGTTGTTTT<br>GGTTTTTCAATTCTTGCTTGTGTTGTTACACGTGAACCATCGGATGATAATG |
| PfSERA2.04 | 10 | TCCAGGTGATATCTTTGTTCCTACGGATCCTCCTGTTCCTGATGATCTTCCTGCTCTTTCGACATCTGATTGGGATACTTCTACACCTGGTCTTGCTGATTC<br>TACTTTTTGTTCTGCTCCTATACCACTTCCTACTTCTTTTTGTTGTTGCTGTTGTCGGGGTTGTGTTTCTTGAGCTAAAGTTGGTAATGCTGGTTGTTCTTTT<br>GGTTTTTCAATTCTTGCTTGTGTTGTTACACGTGAACCATCGGATGATAATG |
| PfSERA2.05 | 9 | TCCAGGTGATATCTTTGTTCCTACGGATCCTCCTGTTCCTGATGATCTTCCTGCTCTTTCGACATCTGATTGGGATACTTCTACACCTGGTTTTGCTAATTC<br>TACTTTTTGTTCTGCTCCTATACCACTTCCTACTTCTTTTTGTTGTTGCTGTTGTTGGGGTTGTGTTTCTTGAGCTAAAGTTGGTAATGCTGGTTGTTGTTTT<br>GGTTTTTCAATTCTTGCTTGTGTTGTTACACGTGAACCATCGGATGATAATG |
| PfSERA2.06 | 8 | TCCAGGTGATATCTTTGTTCCTACGGATCCTCCTGTTCCTGATGATCTTCCTGCTCTTTCGACATCTGATTGGGATACTTCTACACCTGGTCTTGCTGATTC<br>TACTTTTTGTTCTGCTCCTATACCACTTCCTACTTCTTTTTGTTGTTGCTGTTGTTGGGGTTGTGTTTCTTGAGCTAAAGTTGGTAATGCTGGTTGTTGTTTT<br>GGTTTTTCAATTCTTGCTTGTGTTGTTACACGTGAACCATCGGATGATAATG |
| PfSERA2.07 | 7 | TCCAGGTGATATCTTTGTTCCTACGGATCCTCCTGTTCCTGATGATCTTCCTGCTCTTTCGACATCTGATTGGGATACTTCTACACCTGATTTTGCTGATTC<br>TACTTTTTGTTCTGCTCCTATACCACTTCCTACTTCTTTTTGTTGTTGCTGTTGTTGGGGTTGTGTTTCTTGAGCTAAAGTTGGTAATGCTGGTTGTTGTATT<br>GGTTTTTCAATTCTTGCTTGTGTTGTTACACGTGAACCATCGGATGATAATG |
| PfSERA2.08 | 5 | TCCAGGTGATATCTTTGTTCCTACGGATCCTCCTGTTCCTGATGATCTTCCTGCTCTTTCGACATCTGATTGGGATACTTCTACACCTGGTCTTGCTGATTC<br>TACTTTTTGTTCTGCTCCTATACCACTTCCTACTTCTTTTTGTTGTTGCTGTTGTTGGGGTTGTGTTTCTTGAGCTAAAGTTGGTAATGCTGGTTGTTGTATT<br>GGTTTTTCAATTCTTGCTTGTGTTGTTACACGTGAACCATCGGATGATAATG |
| PfSERA2.09 | 5 | TCCAGGTGATATCTTTGTTCCTACGGATCCTCCTGTTCCTGATGATCTTCCTGCTCTTTCGACATCTGATTGGGATACTTCTACACCTGGTCTTGCTGATTC<br>TACTTTTTGTTCTGCTCCTATACCACTTCCTACTTCTTTTTGTTGTTGCTGTTGTCGGGGTTGTGTTTCTTGAGCTAAAGTTGGTAATGCTGGTTGTTCTTTT<br>GGTTCTTCAATTCTTGCTTGTGTTGTTACACGTGAACCATCGGATGATAATG |
| PfSERA2.10 | 4 | TCCAGGTGATATCTTTGTTCCTACGGATCCTCCTGTTCCTGATGATCTTCCTGCTCTTTCGACATCTGATTGGGATACTTCTACACCTGGTATTGCTGATTC<br>TACTTTTTGTTCTGCTCCTATACCACTTCCTACTTCTTTTTGTTGTTGCTGTTGTCGGGGTTGTGTTTCTTGAGCTAAAGTTGGTAATGCTGGTTGTTCTTTT |

| | | |
|---|---|---|
| | | GGTTTTTCAATTCTTGCTTGTGTTGTTACACGTGAACCATCGGATGATAATG |
| PfSERA2.11 | 4 | TCCAGGTGATATCTTTGTTCCTACGGATCCTCCTGTTCCTGATGATCTTCCTGCTCTTTCGACATCTGATTGGGATACTTCTGCACCTGGTTTTGCTGATTC<br>TACTTTTTGTTCTGCTCCTATACCACTTCCTACTTCTTTTTGTTGTTGCTGTTGTTGGGGTTGTGTTTCTTGAGCTAAAGTTGGTAATGTTGGTTGTTGTTTT<br>GGTTTTTCAATTCTTGCTTGTGTTGTTACACGTGAACCATCGGATGATAATG |
| PfSERA2.12 | 3 | TCCAGGTGATATCTTTGTTCCTACGGATCCTCCTGTTCCTGATGATCTTCCTGCTCTTTCGACATCTGATTGGGATACTTCTACACCTGGTTTTGCTGATTC<br>TACTTTTTGTTCTGCTCCTATACCACTTCCTACTTCTTTTTGTTGTTGCTGTTGTTGGGGTTGTGTTTCTTGAGCTAAAGTTGGTAATGCTCGTTGTTGTTTT<br>GGTTTTTCAATTCTTGCTTGTGTTGTTACACGTGAACCATCGGATGATAATG |
| PfSERA2.13 | 3 | TCCAGGTGATATCTTTGTTCCTACGGATCCTCCTGTTCCTGATGATCTTCCTGCTCTTTCGACATCTGATTGGGATACTTCTACACCTGATTTTGCTGATTC<br>TACTTTTTGTTCTGCTCCTATACCACTTCCTACTTCTTTTTGTTGTTGCTGTTGTCGGGGTTGTGTTTCTTGAGCTAAAGTTGGTAATGCTGGTTGTTCTTTT<br>GGTTTTTCAATTCTTGCTTGTGTTGTTACACGTGAACCATCGGATGATAATG |
| PfSERA2.14 | 3 | TCCAGGTGATATCTTTGTTCCTACGGATCCTCCTGTTCCTGATGATCTTCCTGCTCTTTCGACATCTGATTGGGATACTTCTACACCTGGTCTTGCTGATTC<br>TACTTTTTGTTCTGCTCCTATACCACTTCCTACTTCTTTTTGTTGTTGCTGTTGTCGGGGTTGTGTTTCTTGAGCTAAAGTTGGTAATGCTGATTGTTCTTTT<br>GGTTTTTCAATTCTTGCTTGTGTTGTTACACGTGAACCATCGGATGATAATG |
| PfSERA2.15 | 3 | TCCAGGTGATATCTTTGTTCCTACGGATCCTCCTGTTCCTGATGATCTTCCTGCTCTTTCGACATCTGATTGGGATACTTCTACACCTGGTTTTGCTGATTC<br>TACTTTTTGTTCTGCTCCTATACCACTTCCTACTTCTTTTTGTTGTTGCTGTTGTTGGGGTTGTGTTTCTTGAGCTAAAGTTGGTAATGCTGGTTGTTGTTTT<br>TGTTTTTCAATTCTTGCTTGTGTTGTTACACGTGAACCATCGGATGATAATG |
| PfSERA2.16 | 3 | TCCAGGTGATATCTTTGTTCCTACGGATCCTCCTGTTCCTGATGATCTTCCTGCTCTTTCGACATCTGATTGGGATACTTCTGCACCTGGTTTTGCTGATTC<br>TACTTTTTGTTCTGCTCCTATACCACTTCCTACTTCTTTTTGTTGTTGCTGTTGTTGGGGTTGTGTTTCTTGAGCTAAAGTTGGTAATGCTGGTTGTTGTATT<br>GGTTTTTCAATTCTTGCTTGTGTTGTTACACGTGAACCATCGGATGATAATG |
| PfSERA2.17 | 2 | TCCAGGTGATATCTTTGTTCCTACGGATCCTCCTGTTCCTGATGATCTTCCTGCTCTTTCGACATCTGATTGGGATACTTCTGCACCTGGTCTTGCTGATTC<br>TACTTTTTGTTCTGCTCCTATACCACTTCCTACTTCTTTTTGTTGTTGCTGTTGTCGGGGTTGTGTTTCTTGAGCTAAAGTTGGTAATGCTGGTTGTTCTTTT<br>GGTTTTTCAATTCTTGCTTGTGTTGTTACACGTGAACCATCGGATGATAATG |
| PfSERA2.18 | 2 | TCCAGGTGATATCTTTGTTCCTACGGATCCTCCTGTTCCTGATGATCTTCCTGCTCTTTCGACATCTGATTGGGATACTTCTGCATCTGGTTTTGCTGATTC<br>TACTTTTTGTTCTGCTCCTATACCACTTCCTACTTCTTTTTGTTGTTGCTGTTGTTGGGGTTGTGTTTCTTGAGCTAAAGTTGGTAATGCTGGTTGGTCTTT<br>TGGTTTTTCAATTCTTGCTTGTGTTGTTACACGTGAACCATCGGATGATAATG |
| PfSERA2.19 | 2 | TCCAGGTGATATCTTTGTTCCTACGGATCCTCCTGTTCCTGATGATCTTCCTGCTCTTTCGACATCTGATTGGGATACTTCTACACCTGGTTTTGCTGATTC<br>TACTTTTTGTTCTGCTCCTATACCACTTCCTACTTCTTTTTGTTGTTGCTGTTGTTGGGGTTGTGTTTCTTGAGCTAAAGTTGGTAATTCTGGTTGTTGTTTT<br>GGTTTTTCAATTCTTGCTTGTGTTGTTACACGTGAACCATCGGATGATAATG |
| PfSERA2.20 | 2 | TCCAGGTGATATCTTTGTTCCTACGGATCCTCCTGTTCCTGATGATCTTCCTGCTCTTTCGACATCTGATTGGGATACTTCTACACCTGGTTTTGCTAATTC<br>TACTTTTTGTTCTGCTCCTATACCACTTCCTACTTCTTTTTGTTGTTGCTGTTGTCGGGGTTGTGTTTCTTGAGCTAAAGTTGGTAATGCTGGTTGTTCTTTT<br>GGTTTTTCAATTCTTGCTTGTGTTGTTACACGTGAACCATCGGATGATAATG |
| PfSERA2.21 | 2 | TCCAGGTGATATCTTTGTTCCTACGGATCCTCCTGTTCCTGATGATCTTCCTGCTCTTTCGACATCTGATTGGGATACTTCTGCACCTGGTTTTGCTGATTC<br>TACTTTTTGTTCTGCTCCTATACCACTTCCTACTTCTTTTTGTTGTTGCTGTTGTTGGGGTTGTGTTTCTTGAGCTAAAGTTGGTAATGCTGGTTGTTCTTTT<br>GGTTTTTCAATTCTTGCTTGTGTTGTTACACGTGAACCATCGGATGATAATG |
| PfSERA2.22 | 1 | TCCAGGTGATATCTTTGTTCCTACGGATCCTCCTGTTCCTGATGATCTTCCTGCTCTTTCGACATTTGATTGGGATACTTCTACACCTGGTCTTGCTGATTC<br>TACTTTTTGTTCTGCTCCTATACCACTTCCTACTTCTTTTTGTTGTTGCTGTTGTCGGGGTTGTGTTTCTTGAGCTAAAGTTGGTAATGCTGGTTGTTCTTTT |

| | | |
|---|---|---|
| | | GGTTCTTCAATTCTTGCTTGTGTTGTTACACGTGAACCATCGGATGATAATG |
| PfSERA2.23 | 1 | TCCAGGTGATATCTTTGTTCCTACGGATCCTCCTGTTCCTGATGATCTTCCTGCTCTTTCGACATCTGATTGGGATACTTCTACACCTGGTTTTGCTAATTC<br>TACTTTTTGTTCTGCTCCTATACCACTTCCTACTTCTTTTTGTTGTTGCTGTTGTCGGGGTTGTGTTTCTTGAGCTAAAGTTGGTAATGCTGATTGTTCTTTT<br>GGTTTTTCAATTCTTGCTTGTGTTGTTACACGTGAACCATCGGATGATAATG |
| PfSERA2.24 | 1 | TCCAGGTGATATCTTTGTTCCTACGGATCCTCCTGTTCCTGATGATCTTCCTGCTCTTTCGACATCTGATTGGGATACTTCTACACCTGGTCTTGCTGATTC<br>TACTTTTTGTTCTGCTCCTATACCACTTCCTACTTCTTTTTGTTGTTGCTGTTGTTGGGGTTGTGTTTCTTGAGCTAAAGTTGGTAATTCTGGTTGTTGTTTT<br>GGTTTTTCAATTCTTGCTTGTGTTGTTACACGTGAACCATCGGATGATAATG |
| PfSERA2.25 | 1 | TCCAGGTGATATCTTTGTTCCTACGGATCCTCCTGTTCCTGATGATCTTCCTGCTCTTTCGACATCTGATTGGGATCCTTCTACACCTGGTTTTGCTGATTC<br>TACTTTTTGTTCTGCTCCTATACCACTTCCTACTTCTTTTTGTTGTTGCTGTTGTTGGGGTTGTGTTTCTTGAGCTAAAGTTGGTAATGCTGGTTGTTGTTTT<br>GGTTTTTCAATTCTTGCTTGTGTTGTTACACGTGAACCATCGGATGATAATG |
| PfSERA2.26 | 1 | TCCAGGTGATATCTTTGTTCCTACGGATCCTCCTGTTCCTGATGATCTTCCTGCTCTTTCGACATCTGATTGGGATACTTCTACACCTGGTATTGCTGATTC<br>TACTTTTTGTTCTGCTCCTATACCACTTCCTACTTCTTTTTGTTGTTGCTGTTGTTGGGGTTGTGTTTCTTGAGCTAAAGTTGGTAATGCTGGTTGTTGTTTT<br>TGTTTTTCAATTCTTGCTTGTGTTGTTACACGTGAACCATCGGATGATAATG |
| PfSERA2.27 | 1 | TCCAGGTGATATCTTTGTTCCTACGGATCCTCCTGTTCCTGATGATCTTCCTGCTCTTTCGACATCTGATTGGGATACTTCTACACCTGATTTTGCTGATTC<br>TACTTTTTGTTCTGCTCCTATACCACTTCCTACTTCTTTTTGTTGTTGCTGTTGTCGGGGTTGTGTTTCTTGAGCTAAAGTTGGTAATGCTGGTTCTTTTGG<br>TTTTTCAATTCTTGCTTGTGTTGTTACACGTGAACCATCGGATGATAATG |
| PfSERA2.28 | 1 | TCCAGGTGATATCTTTGTTCCTACGGATCCTCCTGTTCCTGATGATCTTCCTGCTCTTTCGACATCTGATTTGGATACTTCTACACCTGATTTTGCTGATTCT<br>ACTTTTTGTTCTGCTCCTATACCACTTCCTACTTCTTTTTGTTGTTGCTGTTGTTGGGGTTGTGTTTCTTGAGCTAAAGTTGGTAATGCTGGTTGTTGTTTT<br>GGTTTTTCAATTCTTGCTTGTGTTGTTACACGTGAACCATCGGATGATAATG |

**Supplemental Table 6. *trap* Haplotypes Detected**

| Haplotype | # Samples Detected | Sequence |
|---|---|---|
| PfTRAP.00 | 3 | TAATCCTCCAGCTATTCCACCTGCAATTTTATATTTATTATCTGATCCTCCTTTTTGTTTATTATTATCTGGCTTTTCATGTTCTTCCCTTTCAGGATGTTTTGGAGTATTGTTATGTTTTCTATTGTATGATCTATTTTCATTATTTCTACCATGTGGACGTGTTTCTCTATCTTCACTATTAGGTACGTGCCTATTTCCATTATTATCTTGACTTTGGGGGTCACTTTGTTTCCTTTCATTATCCAAAACTTTTGGAGGTAATGGTGAATATGGAATATTTCTATCACTTTTATCATTTGGTAAATTATTTTGATT |
| PfTRAP.01 | 2 | TAATCCTCCAGCTATTCCACCTGCAATTTTATATTTATTATCTGATCCTGCTTTTTTTTTATTATTATCTGGCTTTTCATGTTCTTCCCTTTCAGGATGTTTTGGAGTATCGTTATGTTTTCTATTGTATGATCTATTTTCATTATTTCTACCATGTGGACGTGTTTCTCTATCTTCACTATTAGGTACGTGCCTATTTCCATTATTATCTTGACTTTGGGGGTCACTTTGTTTCCTTTCATTATCCAAAACATTTGGAGGTAATGGTGAATATGGAATATATCTATCACTTTTATCATTTGGTAAATTATTTTGATT |
| PfTRAP.02 | 1 | TAATCCTCCAGCTATTCCACCTGCAATTTTATATTTATTATCTGATCCTCCTTTTTTTTTATTATTATCTGGCTTTTCATGTTCTTCCCTTTCAGGATATTTTGGAGTATTGTTATGTTTTCTATTGTATGATCTATTTTCATTATTTCTACCATGTGGACGTGTTTCTCTATCTTCACTATTAGGTACGTGCCTATTTCCATTATTATCTTGACTTTGGGGGTCACTTTGTTTCCTTTCATTATCCAAAACATTTGGAGGTAATGGTGAATATGGAATATATCTATCACTTTTATTATTTGGTAAATTATTTTGATT |
| PfTRAP.03 | 1 | TAATCCTCCAGCTATTCCACCTGCAATTTTATATTTATTATCTGATCCTCCTTTTTTTTTATTATTATCTGGCTTTTCATGTTCTTCCCTTTCAGGATGTTTTGGAGTATCGTTATGTTTTCTATTGTATGATCTATTTTCATTATTTCTACCATGTGGACGTGTTTCTCTATCTTCACTATTAGGTACGTGCCTATTTCCATTATTATCTTGACTTTGGGGGTCACTTTGTTTCCTTTCATTATCCAAAACATTTGGAGGTAATGGTGAATATGGAATATATCTATCACTTTTATCATTTGGTAAATTATTTTGATT |
| PfTRAP.04 | 1 | TAATCCTCCAGCTATTCCACCTGCAATTTTATATTTATTATCTGATCCTCCTTTTTGTTTATTATTATCTGGCTTTTCATGTTCTTCCCTTTCATGATATTTTGGAGTATCGTTATGTTTTCTATTGTATGATCTATTTTCATTATTTCTACCATGTGGATGTGTTTCTCTATCTTCACTATTAGGTACGTGCCTATTTCCATTATTATCTTGACTTTGGGGGTCACTTTGTTTCCTTTCATTATCCAAAACTTTTGGAGGTAATGGTGAATATGGAATATATCTATCACTTTTATCATTTGGTAAATTATTTTGATT |
| PfTRAP.05 | 1 | TAATCCTCCAGCTATTCCACCTGCAATTTTATATTTATTATCTGATCCTCCTTTTTGTTTATTATTATCTGGCTTTTCATGTTCTTCCCTTTCAGGATGTTTTGGAGTATTGTTATGTTTTCTATTGTATGATCTATTTTCATTATTTCTACCATGTGGACGTGTTTCTCTATCTTCACTATTAGGTACGTGCCTATTTCCATTATTATCTTGACTTTGGGGGTCACTTTGTTTCCTTTCATTATCCAAAACATTTGGAGGTAATGGTGAATATGGAATATATCTATCACTTTTATCATTTGGTAAATTATTTTGATT |
| PfTRAP.06 | 1 | TAATCCTCCAGCTATTCCACCTGCAATTTTATATTTATTATCTGATGCTCCTTTTTTTTTATTATTATCTGGCTTTTCATGTTCTTCCCTTTCAGGATATTTTGGAGTATCGTTATATTTTATATTGTATGATCTATTTTCATTATTTCTACCATGTGGACGTGTTTCTCTATCTTCACTATTAGGTACGTGCCTATTTCCATTATTATCTTGACTTTGGGGGTCACTTTGTTTCCTTTCATTATCCAAAACTTTTGGAGGTAATGGTGAATATGGAATATATCTATCACTTTTATCATTTGGTAAATTATTTTGATT |
| PfTRAP.07 | 1 | TAATCCTCCAGCTATTCCACCTGCAATTTTGTATTTATTATCTGATCCTCCTTTTTGTTTATTATTATCTGGCTTTTCATGTTCTTCCCTTTCAGGATGTTTTGGAGTATTGTTATGTTTTCTATTGTATGATCTATTTTCATTATTTCTACCATGTGGACGTGTTTCTCTATCTTCACTATTAGGTACGTGCCTATTTCCATTATTATCTTGACTTTGGGGGTCACTTTGTTTCCTTTCATTATCCAAAACTTTTGGAGGTAATGGTGAATATGGAATATATCTATCACTTTTATCATTTGGTAAATTATTTTGATT |
| PfTRAP.08 | 1 | TAATCCTCCAGCTATTCCACCTGCAATTTTATATTTATTATCTGATCCTCCTTTTTTTTTATTATTATCTGGCTTTTCATGTTCTTCCCTTTCATGATATTTTG |

| | | |
|---|---|---|
| | | GAGTATCGTTATGTTTTCTATTGTATGATCTATTTTCATTATTTCTACCATGTGGATGTGTTTCTCTATCTTCACTATTAGGTACGTGCCTATTTCCATTATT<br>ATCTTGACTTTGGGGGTCACTTTGTTTCCTTTCATTATCCAAAACTTTTGGAGGTAATGGTGAATATGGAATATATCTATCACTTTTATCATTTGGTAAATT<br>ATTTTGATT |
| PfTRAP.09 | 1 | TAATCCTCCAGCTATTCCACCTGCAATTTTATATTTATTATCTGATCCTCCTTTTTGTTTATTATTATCTGGCTTTTCATGTTCTTCCCTTTCATGATATTTTG<br>GAGTATTGTTATGTTTTCTATTGTATGATCTATTTTCATTATTTCTACCATGTGGACGTGTTTCTCTATCTTCACTATTAGGTACGTGCCTATTTCCATTATT<br>ATCTTGACTTTGGGGGTCACTTTGTTTCCTTTCATTATCCAAAACATTTGGAGGTAATGGTGAATATGGAATATATCTATCACTTTTATCATTTGGTAAATT<br>ATTTTGATT |

**Supplemental Table 7. Heterozygosity Contributions.** For each gene, the table shows each haplotype, the number of samples detected, its frequency in the population, and its squared frequency (contribution to homozygosity).

| ***csp*** | | | | ***ama*** | | | | ***sera2*** | | | | ***trap*** | | | |
|---|---|---|---|---|---|---|---|---|---|---|---|---|---|---|---|
| Haplotype | # Samples Detected | Frequency | Contribution_to_Homozygosity | Haplotype | # Samples Detected | Frequency | Contribution_to_Homozygosity | Haplotype | # Samples Detected | Frequency | Contribution_to_Homozygosity | Haplotype | # Samples Detected | Frequency | Contribution_to_Homozygosity |
| PfCSP.00 | 9 | 2.00E-01 | 4.00E-02 | PfAMA1.00 | 23 | 9.75E-02 | 9.50E-03 | PfSERA2.00 | 29 | 1.81E-01 | 3.29E-02 | PfTRAP.00 | 3 | 2.31E-01 | 5.33E-02 |
| PfCSP.01 | 7 | 1.56E-01 | 2.42E-02 | PfAMA1.01 | 18 | 7.63E-02 | 5.82E-03 | PfSERA2.01 | 22 | 1.38E-01 | 1.89E-02 | PfTRAP.01 | 2 | 1.54E-01 | 2.37E-02 |
| PfCSP.02 | 4 | 8.89E-02 | 7.90E-03 | PfAMA1.02 | 17 | 7.20E-02 | 5.19E-03 | PfSERA2.02 | 13 | 8.13E-02 | 6.60E-03 | PfTRAP.02 | 1 | 7.69E-02 | 5.92E-03 |
| PfCSP.03 | 3 | 6.67E-02 | 4.44E-03 | PfAMA1.03 | 16 | 6.78E-02 | 4.60E-03 | PfSERA2.03 | 12 | 7.50E-02 | 5.63E-03 | PfTRAP.03 | 1 | 7.69E-02 | 5.92E-03 |
| PfCSP.04 | 3 | 6.67E-02 | 4.44E-03 | PfAMA1.04 | 15 | 6.36E-02 | 4.04E-03 | PfSERA2.04 | 10 | 6.25E-02 | 3.91E-03 | PfTRAP.04 | 1 | 7.69E-02 | 5.92E-03 |
| PfCSP.05 | 2 | 4.44E-02 | 1.98E-03 | PfAMA1.05 | 12 | 5.08E-02 | 2.59E-03 | PfSERA2.05 | 9 | 5.63E-02 | 3.16E-03 | PfTRAP.05 | 1 | 7.69E-02 | 5.92E-03 |
| PfCSP.06 | 2 | 4.44E-02 | 1.98E-03 | PfAMA1.06 | 11 | 4.66E-02 | 2.17E-03 | PfSERA2.06 | 8 | 5.00E-02 | 2.50E-03 | PfTRAP.06 | 1 | 7.69E-02 | 5.92E-03 |
| PfCSP.07 | 1 | 2.22E-02 | 4.94E-04 | PfAMA1.07 | 11 | 4.66E-02 | 2.17E-03 | PfSERA2.07 | 7 | 4.38E-02 | 1.91E-03 | PfTRAP.07 | 1 | 7.69E-02 | 5.92E-03 |
| PfCSP.08 | 1 | 2.22E-02 | 4.94E-04 | PfAMA1.08 | 11 | 4.66E-02 | 2.17E-03 | PfSERA2.08 | 5 | 3.13E-02 | 9.77E-04 | PfTRAP.08 | 1 | 7.69E-02 | 5.92E-03 |
| PfCSP.09 | 1 | 2.22E-02 | 4.94E-04 | PfAMA1.09 | 10 | 4.24E-02 | 1.80E-03 | PfSERA2.09 | 5 | 3.13E-02 | 9.77E-04 | PfTRAP.09 | 1 | 7.69E-02 | 5.92E-03 |
| PfCSP.10 | 1 | 2.22E-02 | 4.94E-04 | PfAMA1.10 | 9 | 3.81E-02 | 1.45E-03 | PfSERA2.10 | 4 | 2.50E-02 | 6.25E-04 | | | | |
| PfCSP.11 | 1 | 2.22E-02 | 4.94E-04 | PfAMA1.11 | 9 | 3.81E-02 | 1.45E-03 | PfSERA2.11 | 4 | 2.50E-02 | 6.25E-04 | | | | |
| PfCSP.12 | 1 | 2.22E-02 | 4.94E-04 | PfAMA1.12 | 6 | 2.54E-02 | 6.46E-04 | PfSERA2.12 | 3 | 1.88E-02 | 3.52E-04 | | | | |
| PfCSP.13 | 1 | 2.22E-02 | 4.94E-04 | PfAMA1.13 | 6 | 2.54E-02 | 6.46E-04 | PfSERA2.13 | 3 | 1.88E-02 | 3.52E-04 | | | | |

| | | | | | | | | | | | |
|---|---|---|---|---|---|---|---|---|---|---|---|
| PfCSP.14 | 1 | 2.22E-02 | 4.94E-04 | PfAMA 1.14 | 6 | 2.54E-02 | 6.46E-04 | PfSERA 2.14 | 3 | 1.88E-02 | 3.52E-04 |
| PfCSP.15 | 1 | 2.22E-02 | 4.94E-04 | PfAMA 1.15 | 5 | 2.12E-02 | 4.49E-04 | PfSERA 2.15 | 3 | 1.88E-02 | 3.52E-04 |
| PfCSP.16 | 1 | 2.22E-02 | 4.94E-04 | PfAMA 1.16 | 5 | 2.12E-02 | 4.49E-04 | PfSERA 2.16 | 3 | 1.88E-02 | 3.52E-04 |
| PfCSP.17 | 1 | 2.22E-02 | 4.94E-04 | PfAMA 1.17 | 5 | 2.12E-02 | 4.49E-04 | PfSERA 2.17 | 2 | 1.25E-02 | 1.56E-04 |
| PfCSP.18 | 1 | 2.22E-02 | 4.94E-04 | PfAMA 1.18 | 4 | 1.69E-02 | 2.87E-04 | PfSERA 2.18 | 2 | 1.25E-02 | 1.56E-04 |
| PfCSP.19 | 1 | 2.22E-02 | 4.94E-04 | PfAMA 1.19 | 4 | 1.69E-02 | 2.87E-04 | PfSERA 2.19 | 2 | 1.25E-02 | 1.56E-04 |
| PfCSP.20 | 1 | 2.22E-02 | 4.94E-04 | PfAMA 1.20 | 4 | 1.69E-02 | 2.87E-04 | PfSERA 2.20 | 2 | 1.25E-02 | 1.56E-04 |
| PfCSP.21 | 1 | 2.22E-02 | 4.94E-04 | PfAMA 1.21 | 4 | 1.69E-02 | 2.87E-04 | PfSERA 2.21 | 2 | 1.25E-02 | 1.56E-04 |
| | | | | PfAMA 1.22 | 3 | 1.27E-02 | 1.62E-04 | PfSERA 2.22 | 1 | 6.25E-03 | 3.91E-05 |
| | | | | PfAMA 1.23 | 3 | 1.27E-02 | 1.62E-04 | PfSERA 2.23 | 1 | 6.25E-03 | 3.91E-05 |
| | | | | PfAMA 1.24 | 3 | 1.27E-02 | 1.62E-04 | PfSERA 2.24 | 1 | 6.25E-03 | 3.91E-05 |
| | | | | PfAMA 1.25 | 2 | 8.47E-03 | 7.18E-05 | PfSERA 2.25 | 1 | 6.25E-03 | 3.91E-05 |
| | | | | PfAMA 1.26 | 2 | 8.47E-03 | 7.18E-05 | PfSERA 2.26 | 1 | 6.25E-03 | 3.91E-05 |
| | | | | PfAMA 1.27 | 2 | 8.47E-03 | 7.18E-05 | PfSERA 2.27 | 1 | 6.25E-03 | 3.91E-05 |
| | | | | PfAMA 1.28 | 2 | 8.47E-03 | 7.18E-05 | PfSERA 2.28 | 1 | 6.25E-03 | 3.91E-05 |
| | | | | PfAMA 1.29 | 2 | 8.47E-03 | 7.18E-05 | | | | |
| | | | | PfAMA 1.30 | 2 | 8.47E-03 | 7.18E-05 | | | | |

| | | | | | | | |
|---|---|---|---|---|---|---|---|
| | | | | PfAMA 1.31 | 1 | 4.24E-03 | 1.80E-05 |
| | | | | PfAMA 1.32 | 1 | 4.24E-03 | 1.80E-05 |
| | | | | PfAMA 1.33 | 1 | 4.24E-03 | 1.80E-05 |
| | | | | PfAMA 1.34 | 1 | 4.24E-03 | 1.80E-05 |

**Supplemental Table 8. Public *csp* sequences used in analysis**

| Study | Genbank Identifier |
|---|---|
| Foko, LPK, et al. https://doi.org/10.1016/j.gene.2024.148744 | OQ709066.1 |
| | OQ709065.1 |
| | OQ709064.1 |
| | OQ709063.1 |
| | OQ709062.1 |
| | OQ709061.1 |
| | OQ709060.1 |
| | OQ709059.1 |
| | OQ709058.1 |
| | OQ709057.1 |
| | OQ709056.1 |
| | OQ709055.1 |
| | OQ709054.1 |
| | OQ709053.1 |
| | OQ709052.1 |
| | OQ709051.1 |
| | OQ709050.1 |
| | OQ709049.1 |
| | OQ709048.1 |
| | OQ709047.1 |
| | OQ709046.1 |
| | OQ709045.1 |

**Supplemental Figure 1. Amino Acid Frequency in TH2 and TH3 Epitopes.** Frequency determined by presence in invection, not accounting for within sample allele frequency.

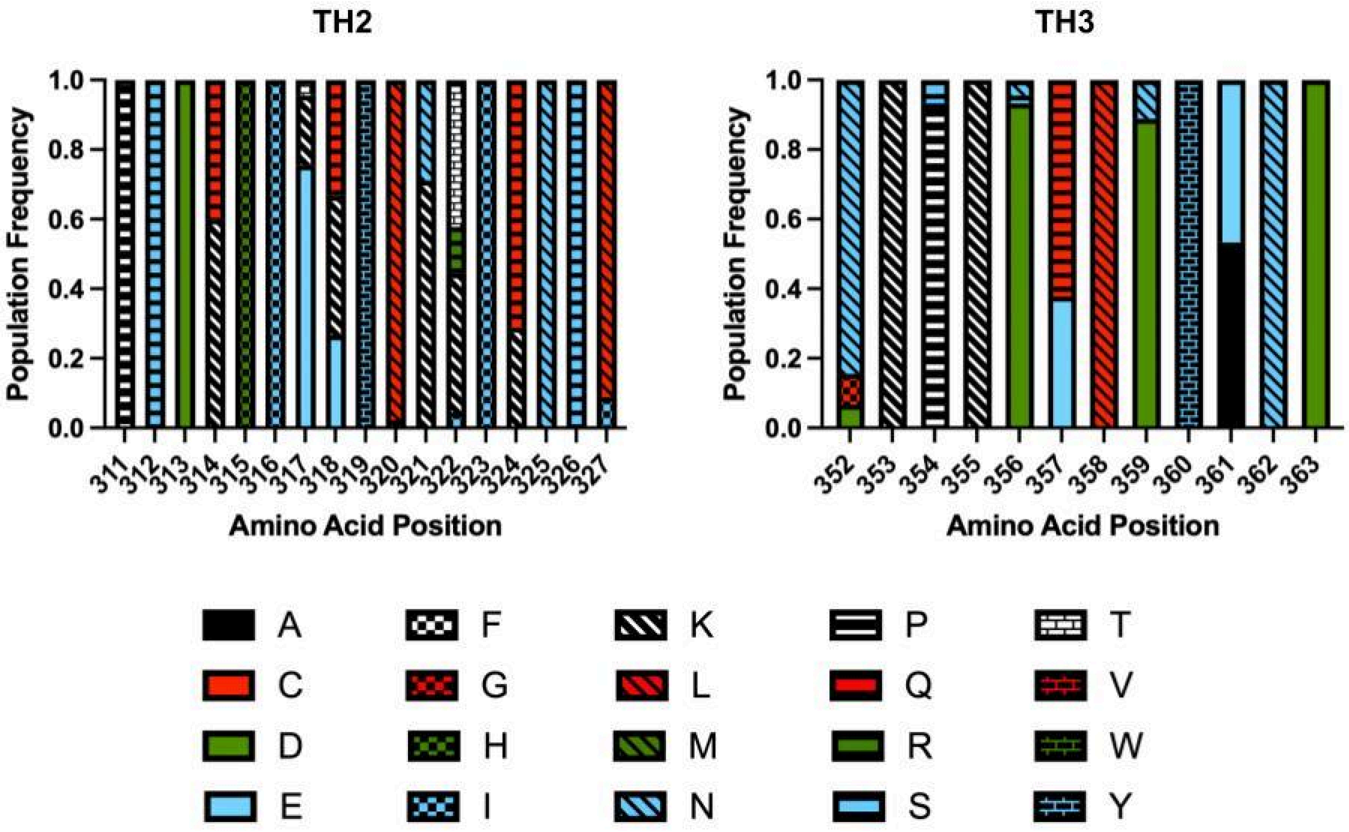

**Supplemental Figure 2. Sliding Window of Tajima's D for *ama* (Panel A), *sera2* (Panel B), and *trap* (Panel C).** Data generated with a window size of 25bp and slide of 5bp.Significant windows are marked with an *.

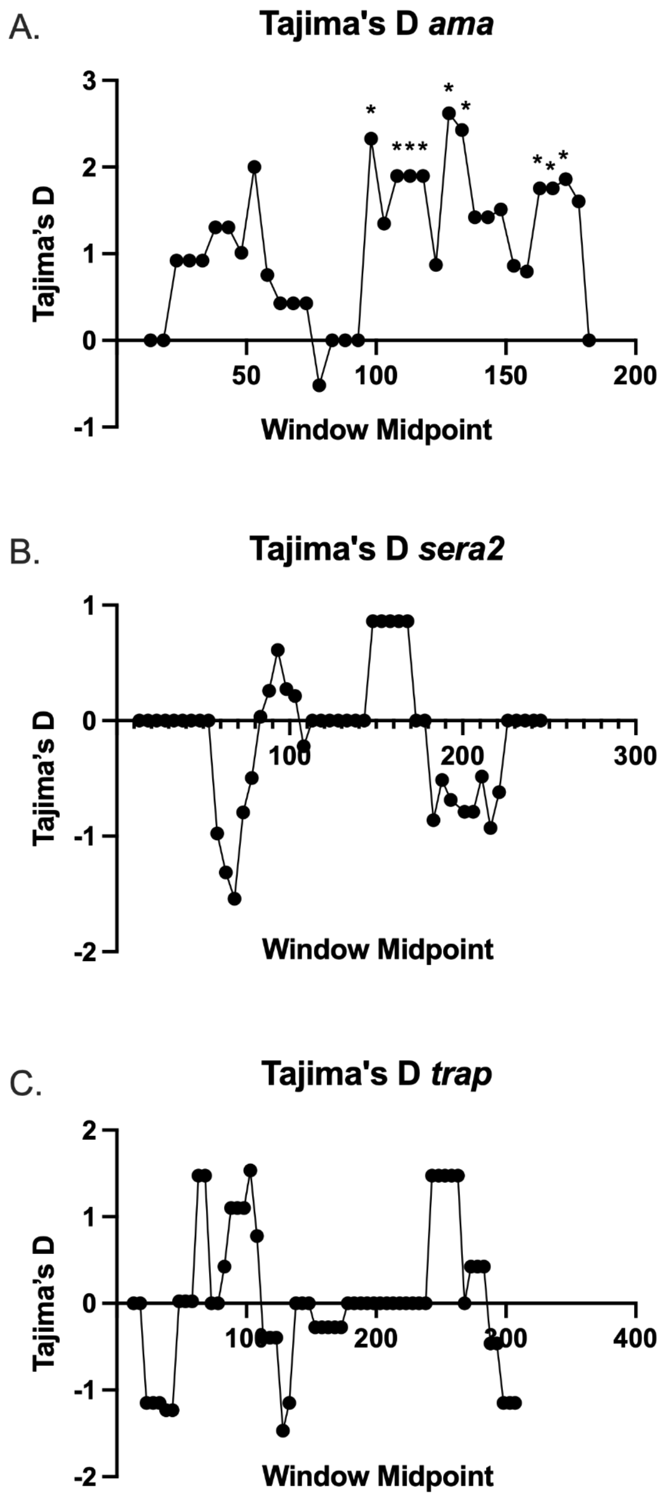
